# Cardiovascular autonomic dysfunction is linked with arterial stiffness across glucose metabolism: The Maastricht Study

**DOI:** 10.1101/2024.12.03.24317865

**Authors:** Jonas R. Schaarup, Lasse Bjerg, Christian S. Hansen, Signe T. Andersen, Marleen van Greevenbroek, Miranda T. Schram, Bastiaan E. de Galan, Coen Stehouwer, Daniel R. Witte

## Abstract

**Objective:** To ascertain the cross-sectional association between cardiovascular autonomic function and arterial stiffness across glucose metabolism status.

**Methods:** We performed a cross-sectional analysis of participants of The Maastricht Study without prior CVD. Cardiovascular autonomic function was based on heart rate variability (HRV) indices from 24-hour electrocardiogram recordings and summarized in Z-scores for time and frequency domains. Aortic and carotid stiffness were assessed by carotid-femoral pulse wave velocity (PWV) and carotid artery distensibility (CD), respectively. We used multiple linear regression to study the associations and adjusted for demographic and lifestyle factors and a range of cardiovascular risk factors. We tested the modification of the associations by glucose metabolism status.

**Results:** PWV and CD measures were available in 3671 and 1806 participants, respectively (median (25^th^; 75^th^ percentile) age: 60 years (53; 66), 51% women, 20 % type 2 diabetes by design. Participants with lower HRV had higher aortic stiffness, as reflected by 2.8% (CI: 2.1; 3.4) and 2.8% (2.1; 3.5) higher PWV per standard deviation (SD) lower composite HRV time-domain and frequency domain Z-score, respectively. Similar trends were observed for carotid stiffness, reflected by 3.2% (1.4; 5.0) and 3.1% (1.2; 5.0) lower CD per SD lower composite HRV time-domain and frequency domain Z-score, respectively. Associations were stronger among people with prediabetes and type 2 diabetes compared to normal glucose metabolism (p-value for interaction for prediabetes: <0.05; and for type 2 diabetes ranging between: <0.05 - <0.10).

**Conclusion:** Cardiovascular autonomic dysfunction is associated with higher aortic and carotid stiffness, especially in people with dysglycemia. Thus, autonomic dysfunction may contribute to cardiovascular risk by affecting vascular stiffness.

**Short abstract:** This study investigates the association between cardiovascular autonomic function and arterial stiffness in 3,671 participants of The Maastricht Study without prior CVD. Cardiovascular autonomic function was assessed using 24-hour heart rate variability (HRV). Aortic stiffness was measured by pulse wave velocity (PWV), and carotid stiffness by carotid artery distensibility (CD). Lower HRV was associated with 2.8–3.2% higher PWV and 3.1–3.2% lower CD per SD decrease in HRV Z-scores, with stronger associations observed in individuals with dysglycemia. These findings suggest that autonomic dysfunction may increase cardiovascular risk through effects on vascular stiffness, particularly in prediabetes and diabetes.

## Background

Improvement of targeted cardiovascular disease (CVD) prevention and treatment in people with diabetes and prediabetes requires a deeper understanding of the interplay between early stages of CVD and diabetes complications [1]. Cardiovascular autonomic dysfunction (autonomic dysfunction), expressed by a reduction in heart rate variability (HRV), is an established risk indicator for CVD that can be easily monitored by wearables, such as smartwatches [2, 3].

However, the mechanisms that explain the link between autonomic dysfunction and CVD remain unclear. Arterial stiffness reflects structural changes in the arterial wall as, with ageing, the elastin fibres gradually are substituted with collagen fibres in the media layer of the large arteries [4]. This remodelling is associated with higher left ventricular afterload contributing to the pathogenesis of heart failure [5, 6]. Moreover, arterial stiffness is linked to atherosclerotic CVD events (e.g. myocardial infarction and stroke) and mortality [7].

Cardiovascular autonomic function can be estimated by HRV indices. The variation between the distance of successive normal RR intervals in milliseconds forms the basic observation underlying all HRV indices. It provides a time- or frequency-domain estimate of the balance between the sympathetic and parasympathetic tone influencing the sinoatrial node [8]. Extended recordings of HRV covering the circadian rhythms of sympathetic and parasympathetic activity may give insight into the role of lower-frequency sources of variability i.e. very low frequency and ultra-low frequency [8]. Lower 24-hour HRV reflects poorer adaptation in cardiac and vascular response to internal and external stimuli throughout the circadian rhythm [9].

Autonomic dysfunction may initially be expressed by sympathetic overactivity and reduced vagal activity[10]. Both in type 1 and type 2 diabetes, autonomic dysfunction and its association with arterial stiffness are well established [12–15]. Moreover, the Whitehall II study showed a longitudinal link in the general population, implying that the association can be observed without the presence of diabetes [16]. However, understanding to what degree the link between autonomic dysfunction and arterial stiffness is modified by dysglycemia is needed to highlight at which stage in the progression of diabetes, autonomic dysfunction is important. Most studies have measured arterial stiffness based on aortic stiffness alone [11]. A separate investigation of both aortic stiffness and carotid stiffness reflects different components of the arterial tree structure that are differently associated with types of CVD events [12, 13].

This etiological cross-sectional study aimed to ascertain the association between cardiovascular autonomic function, measured by 24-hour HRV, and arterial stiffness across glucose metabolism status. We hypothesised that autonomic dysfunction, expressed by lower HRV, is associated with higher levels of aortic and carotid stiffness and that the association is more pronounced in people with more advanced dysglycemia.

## Methods

### Study population

The exact description of The Maastricht Study is referenced from a previous publications [14]: “We used data from The Maastricht Study, an observational prospective population-based cohort study. The rationale and methodology have been described previously. In brief, the study focuses on the aetiology, pathophysiology, complications and comorbidities of type 2 diabetes mellitus (T2DM) and is characterized by an extensive phenotyping approach. Eligible for participation were all individuals aged between 40 and 75 years and living in the southern part of the Netherlands.

Participants were recruited through mass media campaigns and from the municipal registries and the regional Diabetes Patient Registry via mailings. Recruitment was stratified according to known T2DM status, with an oversampling of individuals with T2DM, for reasons of efficiency.

The examinations of each participant were performed within a time window of three months. The study has been approved by the institutional medical ethical committee (NL31329.068.10) and the Minister of Health, Welfare and Sports of the Netherlands (Permit 131088-105234-PG). All participants gave written informed consent. We examined participants who had both HRV and measurements of aortic- and carotid stiffness within a three-month window around the baseline examination round of The Maastricht Study [14].”

The present study includes cross-sectional data from the first 7449 participants, who completed the baseline survey between November 2010 and December 2020 and had measures of arterial stiffness assessed, processed and cleaned. We excluded participants who self-reported prior CVD events, as their pathophysiology and consequent treatment could influence both arterial structural changes and impairment of autonomic balance. We also excluded participants with other types of diabetes than type 2 diabetes, as we investigated the effect modification by glucose metabolism status.

### Exposure

All ECG recordings were obtained using a 12-lead Holter system (Fysiologic ECG Services, Amsterdam, the Netherlands) over 24 hours. The procedure for data collection has previously been reported [15]. During the recording period, participants were instructed to keep their regular daily activities but were asked to refrain from showering. The recorded ECG data were then processed using proprietary Holter Analysis Software at Fysiologic ECG Services (Amsterdam, the Netherlands). Non-sinus cardiac cycles i.e. artefacts and premature/ectopic beats were excluded. This process was subsequently validated through manual inspection. Following the exclusion of non-sinus cardiac cycles, the minimum required recording duration for ECG analysis was set at 18 hours. The software from Fysiologic ECG Services provided the inter-beat intervals in milliseconds (ms) between individual R waves of sinus beats. HRV indices were computed using the publicly available GNU Octave software [16], including the time and frequency domain measures established by the Task Force recommendation on HRV [8]. Time domain HRV indices were calculated, including the standard deviation (SD) of all normal-to-normal (NN) intervals (SDNN, in ms), the SD of the averages of NN intervals in 5-minute segments throughout the recording (SDANN, in ms), the square root of the mean of the sum of squares of differences between adjacent NN intervals (RMSSD, in ms), the mean of the SDs of all NN intervals for all 5-minute segments (SDNN index, in ms), and the NN50 count divided by the total number of all NN intervals (pNN50, percentage). Frequency domain HRV measures were determined using the Fast Fourier Transform based on spectral segment for the whole recording cycle. In the frequency domain HRV, ms² measures the power or energy of the HRV signal within predefined frequency bands.

These included the variance of all NN intervals ≤ 0.4 Hz, total power (TP, in ms^2^), power in the ultralow-frequency range (ULF, in ms^2^ ≤ 0.003 Hz), power in the very-low-frequency range (VLF, in ms^2^; 0.003–0.04 Hz), power in the low-frequency range (LF, in ms^2^; 0.04–0.15 Hz), and power in the high-frequency range (HF, in ms^2^; 0.15–0.4 Hz). We removed outliers in time-domain and frequency HRV indices (see description in the supplementary material). We standardised HRV indices by their mean and SD to make indices comparable and calculated composite z-scores for time and frequency domain HRV indices, respectively. The time-domain Z-score included: SDNN, SDANN, RMSSD, SDNN index, and pNN50, and the frequency-domain Z-score included: TP, HF, LF, VLF, and ULF. Prior evidence shows that this selection of indices covers most of the underlying sources of variance determined by calculations of interbeat intervals [8].

### Outcome

Aortic and carotid stiffness were included as measures for arterial stiffness. The procedure for arterial measurements has been previously documented [17]. Aortic stiffness was determined by carotid-femoral pulse wave velocity (PWV) and was assessed using applanation tonometry (SphygmoCor, Atcor Medical, Sydney, Australia). We included the median value from at least three consecutive PWV recordings in our analyses.

Carotid stiffness was determined by the carotid artery distensibility coefficient (CD). Ultrasound examinations of the left common carotid artery utilizing a 7.5 MHz linear probe-equipped ultrasound scanner (MyLab 70, Esaote Europe, Maastricht, the Netherlands) were conducted to evaluate local carotid distension. Local carotid stiffness was quantified by computing the CD, using the following equation:

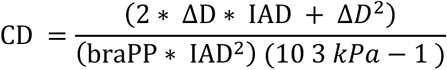

where ΔD represents distension, and braPP signifies brachial pulse pressure. Alongside the vascular assessments, mean heart rate and mean arterial pressure (MAP) were monitored at 5-minute intervals using an oscillometer device (Accutorr Plus, Datascope, Montvale, NJ, USA).

### Covariates

Lifestyle factors of smoking (never, former (quit > 6 months ago), former (quit < 6 months ago), current), physical activity: total (hours/week) and moderate to vigorous exercise (hours/week), and alcohol consumption (average units per week), as well as CVD disease history, and anti-hypertensive, glucose-lowering, and lipid-lowering medication use, were reported through a self-reported questionnaire. Haemoglobin A1c (HbA1c), fasting plasma glucose (FPG), triglycerides, and total, high-density (HDL) and low-density (LDL) cholesterol levels were measured from blood samples. Anthropometric measures of body mass index (BMI) and waist circumference, as well as systolic and diastolic blood pressure, were measured at the study site [14]. We used World Health Organization 2006 criteria for categorizing glucose metabolism status into normal glucose metabolism, prediabetes (impaired fasting glucose and impaired glucose tolerance) and type 2 diabetes, based on a 2-hour 75 gram oral glucose tolerance test (OGTT) and/or the use of glucose lowering medication [18]. HbA1c was not used as criterion for type 2 diabetes or prediabetes.

### Statistical analysis

We describe population characteristics by the distribution (median, 25^th^ and 75^th^ percentile) for continuous variables and frequencies (numbers, percentage) for categorical variables.

We performed multiple linear regression with heart rate variability indices as exposure for the outcome of arterial stiffness. We included the glucose metabolism status (normal glucose metabolism, prediabetes, and type 2 diabetes) to account for the oversampling of individuals with known type 2 diabetes. We further adjusted for mean arterial pressure (MAP) to account for potential instrumental bias, ensuring that elevated MAP during the measurement of arterial stiffness does not falsely indicate greater stiffness [19]. Model 1 was adjusted for age, sex, education, MAP, and diabetes status. In model 2 we further adjusted for self-reported total physical activity (hours/week), smoking behaviour, alcohol use, body mass index, HbA1c, triglycerides, total-to-high density lipoprotein cholesterol ratio, lipid-modifying- and antihypertensive medication. Blood pressure measures other than MAP are considered a collider as they are affected by autonomic dysfunction and arterial stiffness and thus were not included in the model [20]. To obtain normally distributed residuals, we log-transformed measures for arterial stiffness (PWV and CD) and back-transformed the model estimates into a percentage scale. We further tested for effect modification by sex and diabetes status by including them as multiplicative interaction terms, in separate models. A significant interaction was determined by a p-value<0.05. We also carried out a subsidiary analysis to investigate possible gradual stratified modification by higher glucose levels, using 20th percentiles of either FPG or HbA1c after excluding people using glucose-lowering medication. To test the robustness of our analysis we performed a sensitivity analysis first excluding individuals with antihypertensive treatment and subsequently people with type 2 diabetes. In the effect modification analysis by diabetes status, we performed an additional analysis excluding people using betablockers. We performed a complete case analysis, using the statistical program R (4.3.2) [21].

## Results

### Descriptive

Of the whole study population with available measures of HRV without prior CVD events and other types of diabetes, 3671 had PWV and 1806 had CD measured. Fifty-one percent were women and participants had a median (25^th^; 75^th^ percentile) age of 60 (53; 66) years, and 2,387 (65%), 537 (15%), and 747 (20%) had normal glucose metabolism, prediabetes, and type 2 diabetes, respectively. The population with type 2 diabetes more frequently used lipid-lowering and anti-hypertensive medication compared to the populations with prediabetes or normal glucose metabolism (see supplementary table 3S). The median SDNN (HRV) was 133 ms (110; 158). The median PWV (Aortic stiffness) was 8.40 (7.44; 9.76) m/s and CD (Carotid stiffness) 14.1 (11.0; 17.8) 10^-3^/kPa.

### Heart rate variability and aortic stiffness

In model 1, for each SD lower HRV time-domain Z-score, PWV was 2.78% (CI: (2.13; 3.42) higher. For each SD lower HRV frequency-domain Z-score, PWV was 2.82% (CI: 2.14; 3.49) higher (see Fig. 3A and 3B). The strongest associations were seen in SDNN and SDANN for the time domain and in total power, VLF, and ULF for the frequency domain (see Fig. 2A). Associations did not change materially upon adjustment for the confounders in model 2. The sensitivity analyses showed that excluding participants using antihypertensive medication did not materially change the estimates. No interaction was observed by sex (see supplementary material: table 8S).

**Figure 1:**
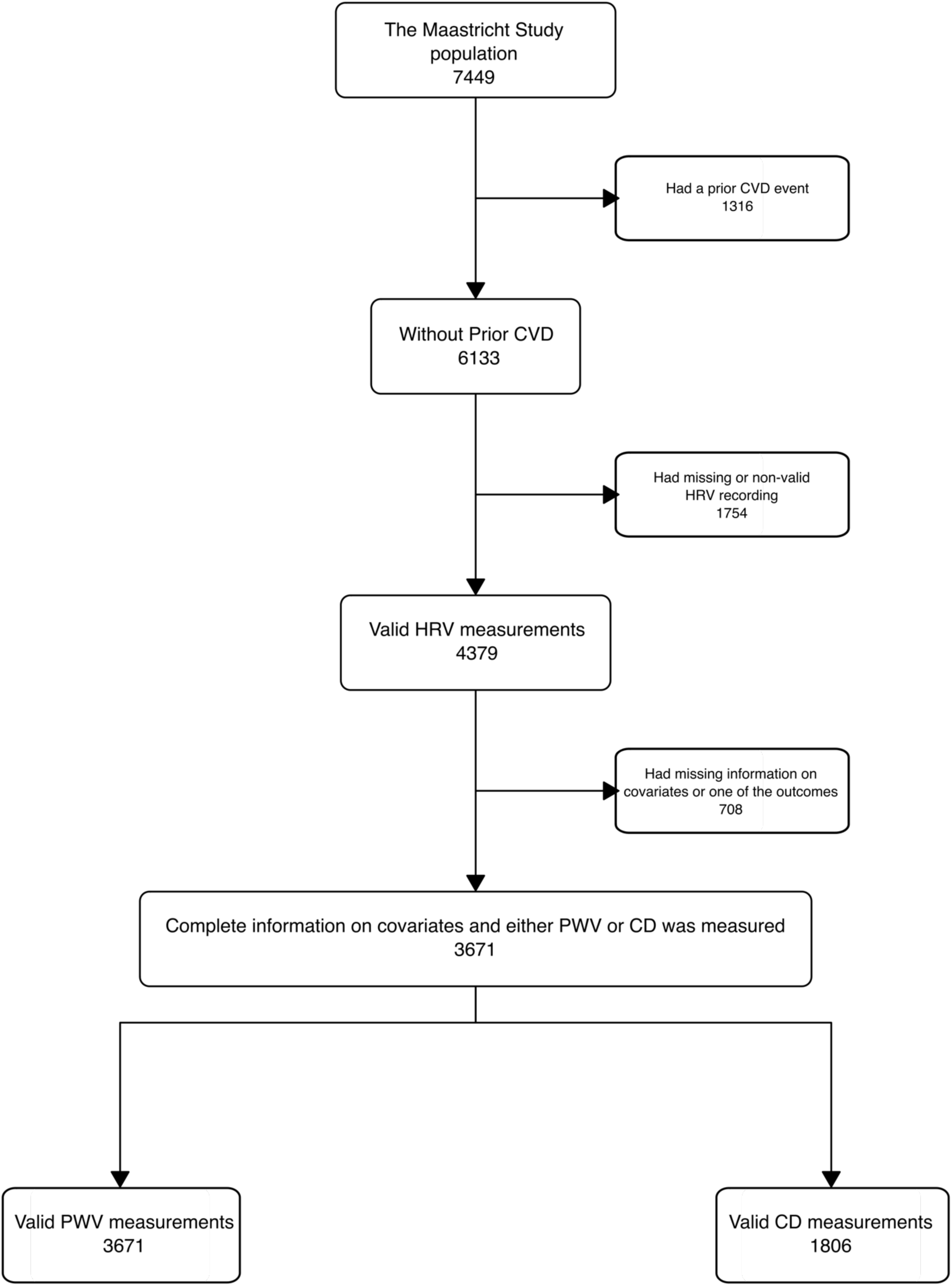
Study flowchart.

**Table 1:** Study population characteristics.

| Characteristic | N = 3,671 |
| --- | --- |
| Sex |  |
| Men | 1,772 (48%) |
| Women | 1,899 (52%) |
| Age (years) | 60 (53, 66) |
| Ethnicity |  |
| White | 3,630 (99%) |
| Non-white | 41 (1.1%) |
| Education |  |
| Low (No education, (un)completed primary education, or lower vocational education) | 1,097 (30%) |
| Middle (intermediate vocational education or higher secondary education) | 1,050 (29%) |
| High (Higher vocational education or university education) | 1,524 (42%) |
| Alcohol consumption |  |
| None | 606 (17%) |
| Low (Women: $\leq 7$, Men: $\leq 14$) | 2,149 (59%) |
| High (Women: $> 7$, Men: $> 14$) | 916 (25%) |
| Alcohol total (g/day) | 9 (2, 19) |
| Smoking status |  |
| Never | 1,414 (39%) |
| Former (quit $> 6$ months ago) | 1,734 (47%) |
| Former (quit $< 6$ months ago) | 62 (1.7%) |
| Current | 461 (13%) |
| Total physical activity (hours/week) | 13 (8, 19) |
| Moderate to vigorous physical activity (hours/week) | 4.5 (2.3, 7.8) |
| BMI ($\text{kg}^2/\text{m}$) | 26.0 (23.6, 28.8) |
| Waist (cm) | 93 (85, 102) |
| HbA1c (mmol/mol) | 37 (34, 41) |
| HbA1c (%) | 5.54 (5.26, 5.90) |
| Fasting plasma glucose (mmol/L) | 5.40 (4.90, 6.00) |
| LDL (mmol/L) | 3.10 (2.40, 3.80) |
| HDL (mmol/L) | 1.50 (1.20, 1.90) |
| Total cholesterol (mmol/L) | 5.30 (4.60, 6.10) |
| Triglycerides (mmol/L) | 1.18 (0.87, 1.65) |
| Total cholesterol-to-HDL cholesterol ratio | 3.40 (2.77, 4.25) |
| Glucose metabolism status |  |
| Normal glucose metabolism | 2,387 (65%) |
| Prediabetes | 537 (15%) |
| Type 2 Diabetes | 747 (20%) |
| Duration of type-2 diabetes (only for diagnosed participants) | 3 (0, 9) |
| Mean IBI (ms) | 828 (765, 903) |
| SDNN (ms) | 133 (110, 158) |
| RMSSD (ms) | 25 (20, 34) |
| SDANN (ms) | 119 (97, 144) |
| SDNNi (ms) | 52 (42, 63) |
| pNN50 (%) | 6 (3, 12) |
| TP (ms<sup>2</sup>) | 11,547 (7,986, 16,385) |
| ULF (ms<sup>2</sup>) | 9,785 (6,653, 14,190) |
| VLF (ms<sup>2</sup>) | 1,102 (736, 1,566) |
| LF (ms<sup>2</sup>) | 364 (221, 590) |
| HF (ms<sup>2</sup>) | 84 (50, 149) |
| Systolic blood pressure (mmHg) | 126 (116, 136) |
| Diastolic blood pressure (mmHg) | 76 (71, 81) |
| Mean arterial pressure (mmHg) | 96 (89, 103) |
| Carotid artery distensibility ($10^{-3}$/kPa) | 14.1 (11.0, 17.8) |
| Carotid-femoral pulse wave velocity (m/s) | 8.40 (7.44, 9.76) |
| Diagnosed hypertension | 1,741 (47%) |
| Glucose-lowering medication | 518 (14%) |
| Antihypertensive medication | 1,110 (30%) |
| Beta blockers | 420 (11%) |
| Diuretic aldosterone | 15 (0.4%) |
| Diuretics | 471 (13%) |
| Lipid-lowering medication | 906 (25%) |

Data are shown as n (%) or median (IQR)

**Figure 2:**
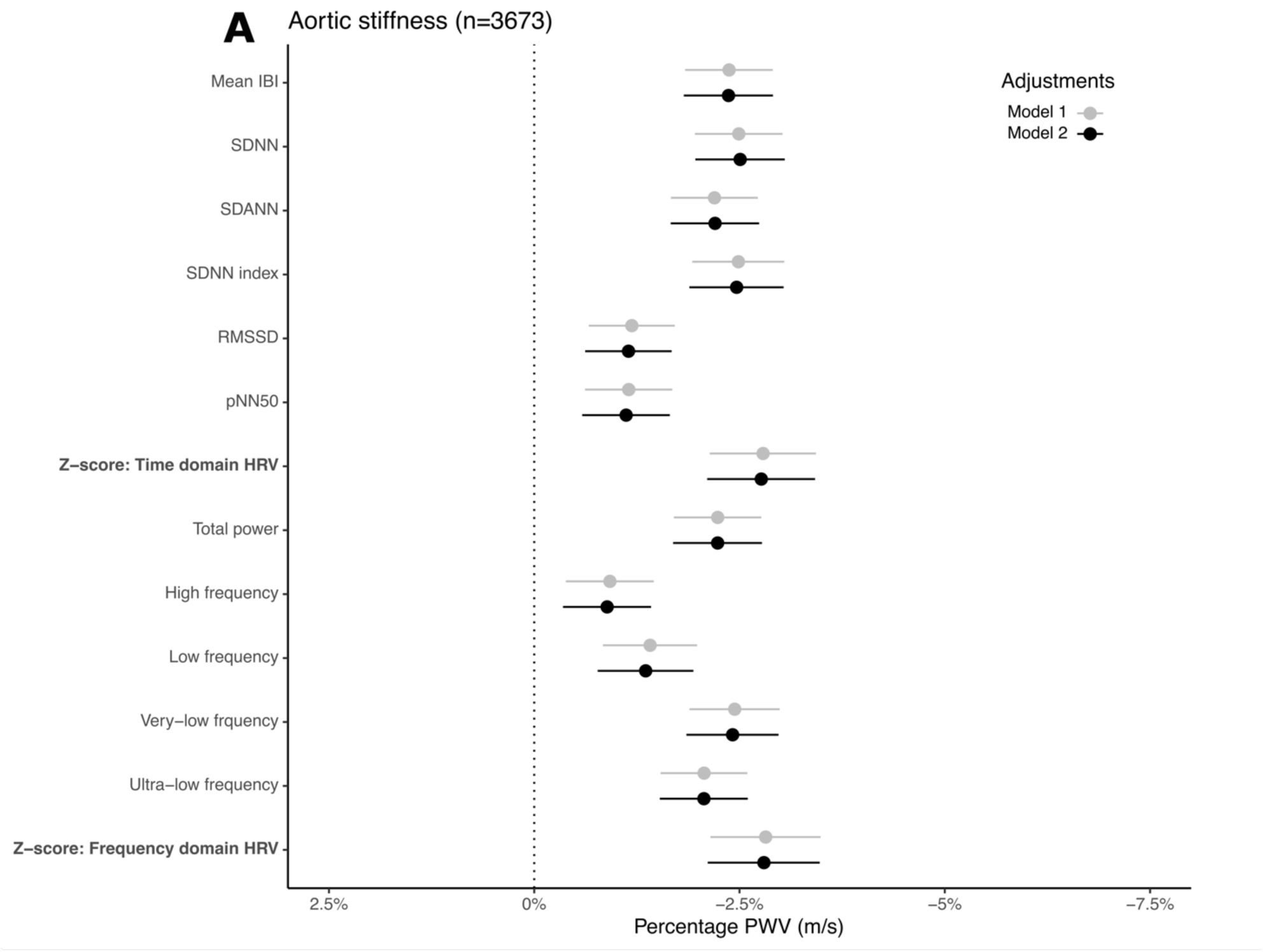
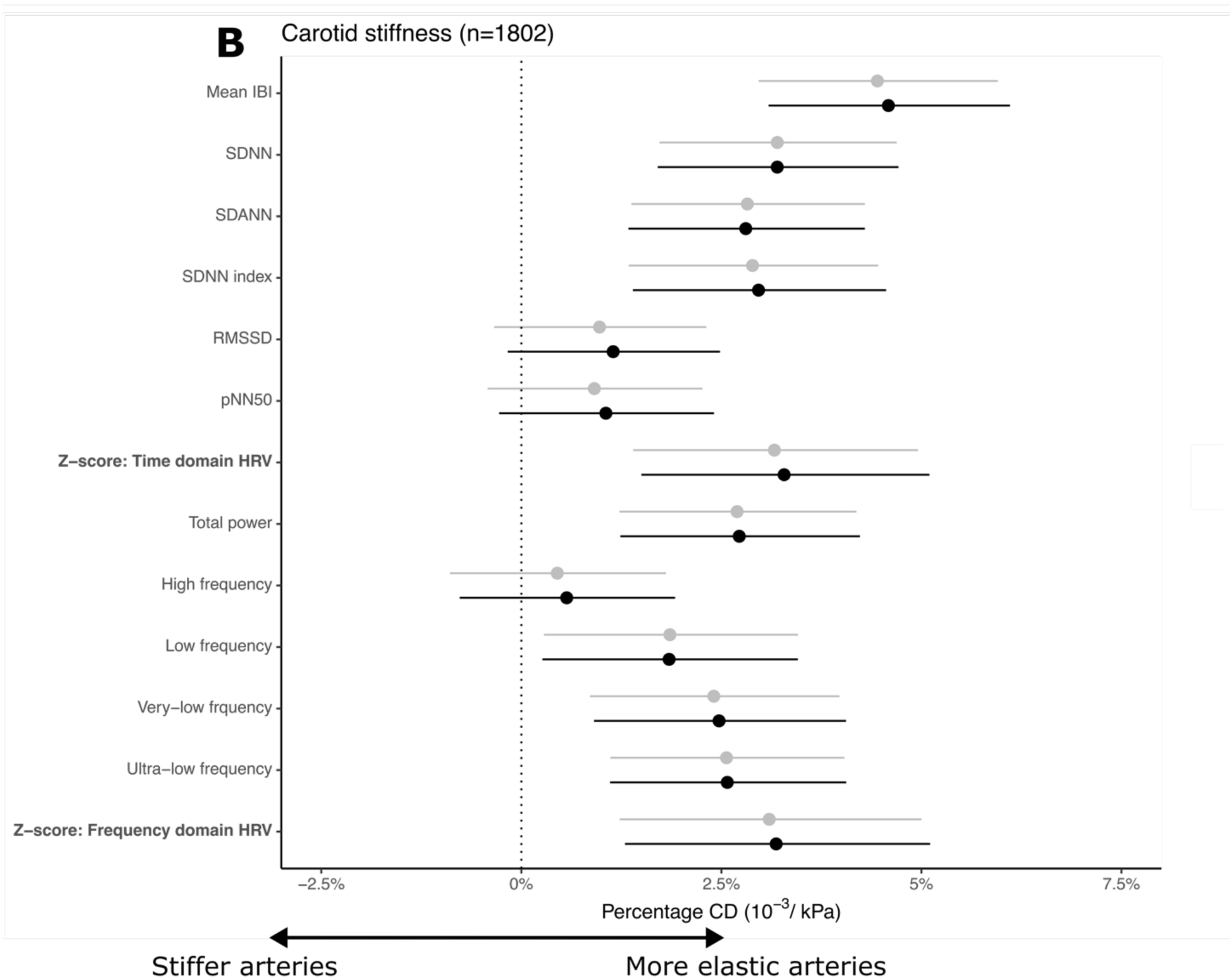
Association between long-term HRV and arterial stiffness. Percentage PWV (**A**) or CD (**B)** per SD increase in heart rate variability index and heart period intervals. Model 1: adjusted for sex, age, educational status, diabetes status, and mean arterial pressure. Model 2: Model 1 + physical activity, smoking behaviour, alcohol use, body mass index, hba1c, triglycerides, total-to-high density lipoprotein cholesterol ratio, lipid-modifying- and antihypertensive medication. Z-score: Frequency domain HRV

**Figure 3:**
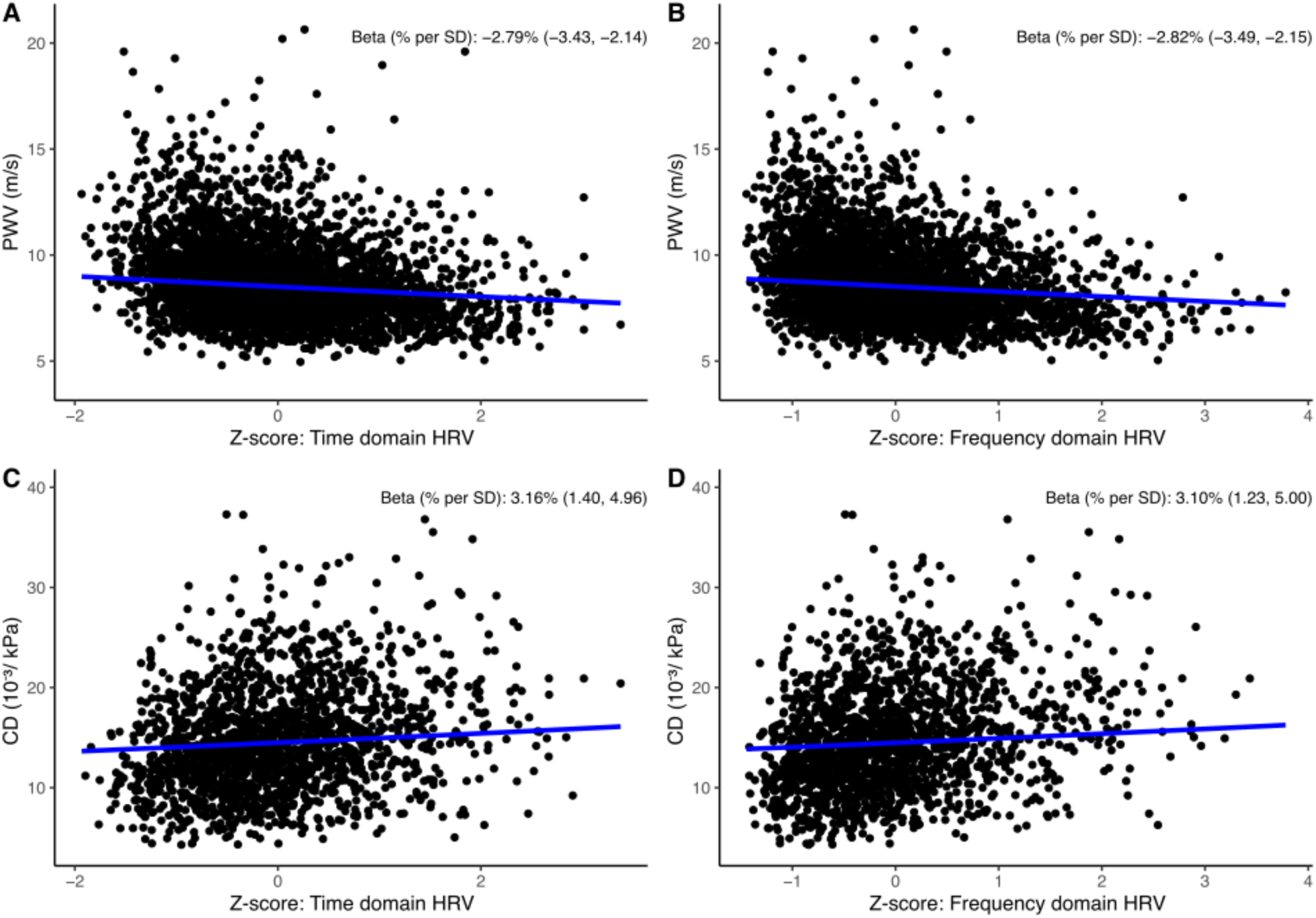
Linear relationship between HRV and aortic and carotid stiffness. **A:** Percentage PWV per SD in time-domain composite z-score **B:** Percentage PWV per SD in frequency-domain composite z-score **C:** Percentage higher CD per SD in time-domain composite z-score **D:** Percentage CD per SD in frequency-domain composite z-score. All regression lines were adjusted for being a male, 60 years old, low educational level, without prediabetes or type-2 diabetes, and with 96mmHg mean arterial pressure.

### Heart rate variability and carotid stiffness

In model 1, for each SD lower HRV time-domain Z-score, CD was 3.17% (CI: 1.41; 4.96) lower. For each SD lower HRV frequency-domain Z-score, CD was 3.12% (CI: 1.24; 5.01) lower (see Fig. 3C and 3D). The strongest associations were seen in SDNN and SDANN for time-domain indices and in total power, VLF, and ULF for the frequency domain (see Fig. 2AB). Associations did not change materially upon adjustment for the confounders in model 2. Except for HRV index VLF, the sensitivity analyses showed that excluding participants using antihypertensive medication did not materially change the estimates. No interaction was observed by sex (see supplementary material: table 9S).

### Effect modification by glucose metabolism

The association between HRV and measures of arterial stiffness was stronger in people with prediabetes and type 2 diabetes than in those with normal glucose metabolism (see Fig. 4AB). Indeed, we observed statistically significant interactions when comparing prediabetes and with normal glucose metabolism, whereas the interaction was only significant for type 2 diabetes in the association between HRV frequency-domain Z-score and PWV. Excluding people using betablockers raised the estimates for the type 2 diabetes group in the analysis with PWV as outcome but not in CD (see Fig 3S). Effect modification estimates for each HRV index are presented in the supplementary material (see Table 6S and 7S). When we excluded people using glucose lowering-medication and analysed the stratified modification by quintiles of glycaemia, we found stronger associations between the frequency and time-domain Z-score and PWV and CD in higher percentiles of FPG and HbA1c (see supplementary material Fig1S and Fig2S).

**Figure 4:**
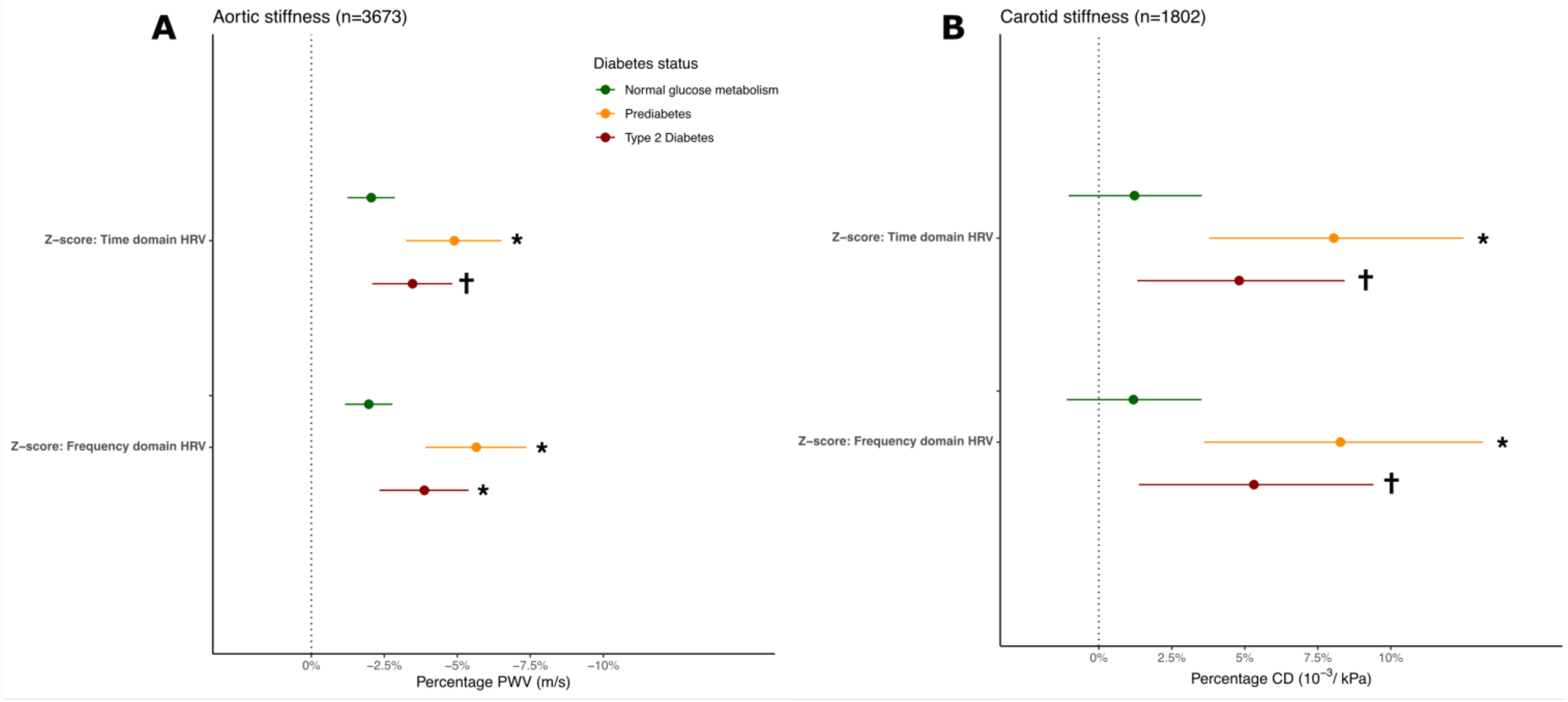
Association between long-term HRV and arterial stiffness modified by diabetes status. **A:** Percentage PWV per SD in time-domain and frequency-domain composite z-score by diabetes status **B:** Percentage CD per SD in time-domain and frequency-domain composite z-score by diabetes status. Estimates are adjusted for sex, age, educational status, mean arterial pressure, physical activity, smoking behaviour, alcohol use, body mass index, HbA1c, triglycerides, total-to-high density lipoprotein cholesterol ratio, lipid-modifying- and antihypertensive medication. Normal glucose metabolism was defined as reference group. *Interaction term p-value < 0.05 ^┼^Interaction term p-value < 0.10

## Discussion

In this study, we showed that cardiovascular autonomic dysfunction, determined by long-term HRV, was associated with both aortic and carotid stiffness among older adults, irrespective of the presence of glucose metabolism status, although the association was stronger in those with prediabetes or type 2 diabetes. Lower HRV was associated with higher stiffness either measured by higher PWV or lower CD.

The association for the time domain Z-score was mainly driven by SDNN and SDANN and that for the frequency domain Z-score was primarily driven by total power, VLF, and ULF. Hence, the associations were mostly determined by HRV indices calculated by global variation of interbeat intervals and lower frequency bands capturing long oscillations in interbeat intervals. Across the HRV indices, SDNN was most strongly associated with measures of arterial stiffness.

The magnitude of the observed associations was modest but relevant when compared to equivalent associations of age with arterial stiffness. One SD lower HRV was equivalent to the effect of 2.7 additional years on PWV and to 1.6 years for CD. A hypothetical individual (male, non-smoker, low alcohol consumption, no diabetes or hypertension, and mean values for all continuous confounders in model 2) with an SDNN at the 10^th^ percentile (90 ms) had 8.77 m/s PWV, and 14.3 10^-3^/kPa CD. A similar hypothetical individual at the 90^th^ percentile of SDNN (180 ms) had a 6.5 % lower PWV and 8.2 % higher CD. Hence, lower 24-hour HRV is associated with arterial stiffness characterized both by local stiffness at the carotid site and by dynamic alterations in the aorta. The link between long-term HRV and these distinctly measured surrogate CVD markers, suggests that long-term HRV is likely also linked to the risk of ischemic and stroke events. Earlier studies have shown that short-term HRV is linked with both coronary heart disease and stroke but are less conclusive with regard to long-term HRV [22, 23].

We accounted for the oversampling of people with type 2 diabetes by adjusting for diabetes status and correcting for the instrument bias in stiffness measures, caused by higher pulse pressure during measurement of PWV and CD, by adjusting for MAP. Adjustment for lifestyle habits and cardiovascular risk factors did not materially change the estimates, suggesting most of the measurable confounding was captured by diabetes status and MAP. In our sensitivity analysis, without participants on anti-hypertensive treatment, the estimates did not materially change, thus we focused on models for the entire study population and adjusted for medication in the full model.

Several studies found lower HRV indices to be associated with aortic stiffness among people with either type 1 or type 2 diabetes [11, 24]. Our study extends these findings by showing that the associations are already present in people without diabetes, albeit to a lesser degree than in people with prediabetes or diabetes.

Both cardiovascular autonomic dysfunction and arterial stiffness are likely to be shared consequences of cardiometabolically disturbed environment including dyslipidaemia, hyperinsulinemia and advanced glycation end-products induced by hyperglycaemia, oxidative stress, and inflammation [15, 25–27] [28, 29]. Our results support the notion that hyperglycaemia modifies the association between HRV and arterial stiffness, as we found stronger associations in the higher quintiles of both FPG and HbA1c in participants without glucose-lowering medication. Early deterioration of glucose metabolism starts a complex cycle of complications, in this case, autonomic dysfunction that along with dysglycemia, likely through neuronal damage[15], contributes to vascular dysfunction. Although our data is cross-sectional, the effect modification gives a notion that the CVD risks are higher in prediabetes and improvement of glycaemic control could potentially in part modify the contribution of low HRV to arterial stiffness.

Two explanations might clarify why the effect modification did not increase progressively by glycaemic status in the present study. First, because of being diagnosed with type 2 diabetes, participants were more likely to receive cardioprotective care, including glucose-lowering, lipid-lowering, and antihypertensive medication, an effect that cannot be accounted for by adjustment. After exclusion of people using betablockers, the results partly explained why type 2 diabetes showed a smaller modifying effect compared to prediabetes in the outcome of PWV, but not in the outcome of CD. The second explanation could be due to selection bias, as participants with type 2 diabetes, who participated in the Maastricht Study and underwent both long-term ECG recordings and measures of arterial stiffness might be healthier than the background population with type 2 diabetes.

We showed that shorter mean IBI was associated with both aortic and carotid stiffness, emphasising a potential mediating role of higher heart rate in autonomic dysfunction. Sympathetic predominance may result in a higher heart rate and hence lead to higher shear stress on the arterial wall. The association might also be driven by direct sympathetic effects on arteries, caused by increased levels of norepinephrine and reduced clearance [30, 31]. We cannot exclude that the association between HRV and arterial stiffness might be bidirectional, hence arterial remodelling may also cause changes in autonomic balance, which might particularly be expressed in carotid stiffness. The baroreflex receptors located in the carotid artery region become less sensitive as compliance in the carotid region deteriorates, which may result in less adaptive heart rate and blood pressure response [32, 33].

In summary, cardiovascular autonomic function might be a relevant risk indicator of efforts to prevent trajectories towards CVD mediated through arterial stiffness, even before the onset of diabetes. Our findings support the view that lower 24-hour HRV is an indicator of elevated CVD risk.

Hyperglycaemia is rarely an isolated risk factor among people with prediabetes and type 2 diabetes. Therefore, current type 2 diabetes guidelines focus on multifactorial cardiometabolic management, the effect of which on micro- and macrovascular complications has been clearly demonstrated [10, 34]. Closer attention to the mechanisms that mediate these effects offers the prospect of new intervention points. Although it is conceivable that multifactorial risk management slows the progression of arterial stiffening partially by modulating autonomic dysfunction, it remains to be proven whether modification of HRV per se contributes causally to reduction of CVD risk. To ascertain this causality, observational studies using Mendelian randomisation would provide a first line of evidence. Furthermore, cardiometabolic trials, assessing either lifestyle modification or pharmacological intervention should, if possible, measure HRV to enable a structured mediation analysis.

Our findings help us understand that the progression of autonomic dysfunction plays a role in CVD risk and confirm that prediabetes defines a group with a higher risk of complications. Lifestyle and glucose-lowering interventions improve cardiometabolic outcomes in prediabetes but have not yet been shown to effectively prevent CVD or all-cause mortality events [35]. Autonomic dysfunction may serve as a tool for risk stratification among individuals with prediabetes who have high CVD risk. These individuals may particularly benefit from lifestyle interventions to reduce their CVD risk [36, 37]. Lastly, our findings show that autonomic dysfunction plays a smaller, but still meaningful, role in CVD risk among people without diabetes.

The strengths of the study are the large sample size with a large subpopulation with type 2 diabetes and that HRV was determined by long-term 24-hour ECG recordings in free-living conditions. Recordings of 24-hour ECG traces provide a full day measurement of cardiovascular autonomic function during the circadian rhythm, including responses in free living conditions [8]. There are also limitations to consider. First, during ECG recordings, non-stationary activity (including physical activity, meals, consumption of caffeine.) might influence the assessment of cardiovascular autonomic function [8]. Second, the level of HRV may depend on heart rate. We did not include adjustment of heart rate in the model as we believe it violates the principles of multicollinearity. Moreover, as a higher heart rate is determined by increased sympathetic bursts, we consider it to be a mediator on the pathway to arterial stiffness [31]. We have a full-day recording capturing heartbeats in rest and activity. These measures are representative of valid autonomic assessment in a full-day cycle [8]. We believe it is more relevant to consider the correction for heart rate in short-term recordings, as random factors (e.g. time of the day, smoking, caffeine intake) can influence this standardized recording procedure [34]. Therefore, we argue that, in the current study, heart rate should not be included as an adjustment for either confounding or instrumental bias. Third, the generalizability of our findings is limited to populations including middle-aged white people with access to high-quality diabetes care. Finally, our study is based on cross-sectional data and thus, we cannot infer a causal direction. However, we attempted to mimic an aetiological ordering by showing the temporality of glucose metabolism (normal, prediabetes, and type 2 diabetes) in the relationship between autonomic dysfunction and arterial stiffness.

Longitudinal data from the Whitehall study showed that a steeper decrease in short-term 5-min HRV over 10 year was associated with subsequent higher levels of aortic stiffness in a five-year trajectory [24]. This suggests that autonomic dysfunction is mainly contributing to arterial stiffness rather than the other way around.

Higher physical activity is longitudinally associated with increased HRV [35]. We included self-reported total physical activity to account for habitual physical activity, but we did not include accelerometer-based physical activity adjustment as this might result in over-adjusting for the concurrent physical movement on the concurrent day of the HRV recordings. Earlier data from The Maastricht Study confirmed that adjustment for objective physical activity by mean stepping time measured by an accelerometer did not change the estimates of their analysis of HRV compared to self-reported physical activity [15].

Wearable devices have made data collection of physiological measures (e.g. pulse rate, blood oxygen saturation, physical activity etc.) more accessible in general populations e.g. by smartwatches. Global distributed HRV measures as well as lower frequency bands have been shown to be valid [2]. Hence, long-term HRV is becoming more accessible to users and eventually health care providers, however its clinical relevance and role remain to be ascertained before implementation. Our study shows that lower HRV is associated with surrogate markers for CVD risk, even at normal glucose metabolism. Thus, a cycle of 24-hour long-term HRV measured by wearable devices might be an easy and non-invasive tool to detect people who silently have higher CVD risk in all stages of glucose metabolism, beyond conventional CVD markers.

## Conclusion

Lower 24-hour HRV was associated with both higher aortic and carotid stiffness. This association was stronger with worse glucose metabolism status. Cardiovascular autonomic dysfunction may contribute to cardiovascular risk by affecting vascular stiffness. The prognostic value of 24-hour HRV in CVD risk, and whether the CVD risk reduction of glucose-lowering intervention is mediated by improved cardiovascular autonomic function remain open for further investigation.

## Data Availability

The data of this study derive from The Maastricht Study, but restrictions apply to the availability of these data, which were used under license for the current study. Data are, however, available from the authors upon reasonable request and with permission of The Maastricht Study management team.

## Abbreviations

CAN: Cardiovascular autonomic neuropathy
CD: Carotid artery distensibility
PWV: carotid-femoral pulse wave velocity
MAP: Mean arterial pressure
CVD: Cardiovascular disease
ECG: Electrocardiogram
HRV: Heart rate variability
rHR: Resting heart rate
SDNN: The standard deviation of normal-to-normal R-R intervals
SDANN: The standard deviation of the averages of NN intervals in 5-minute segments throughout the recording
SDNN index: The mean of the standard deviation of all NN intervals for all 5-minute segments
pNN50: The NN50 count divided by the total number of all NN intervals
RMSSD: The square root of the mean of the sum of squares of differences between adjacent NN intervals
TP: Total frequency
HF: High frequency
LF: Low frequency
VLF: Very-low frequency
ULF: Ultralow frequency

## Acknowledgements

We want to acknowledge all participating women and men in the Maastricht study. We would like to thank Tan Lai Zhou for his expertise in The Maastricht Study of the heart rate variability recordings. This study has been part of a collaboration starting from the European Association of Studies in Diabetes (EASD) Scientist training program 2022 in Maastricht.

## Authors’ contributions

JRS, LB, and DW contributed to the conception and design of the study, performed data analysis and interpretation, drafted the manuscript, critically revised it for significant intellectual content, and gave final approval of the version to be published. CS contributed to the conception and design, assisted with data acquisition, interpreted the data, critically revised the manuscript for important intellectual content, and approved the final version for publication. CSH and STA contributed to the conception and design, participated in data analysis and interpretation, critically revised the manuscript for intellectual content, and approved the final version for publication.

MvG, MTS, and BEdG were involved in data acquisition, critically revised the manuscript for significant intellectual content, and approved the final version for publication. JRS is the guarantor of this work, having full access to all study data, and is responsible for the integrity and accuracy of the data analysis.

## Ethics

The study has been approved by the institutional medical ethics committee (NL31329.068.10) and the Minister of Health, Welfare and Sports of the Netherlands (Permit 131088-105234-PG). All participants gave written informed consent.

