## Supplementary material for "Cardiovascular autonomic dysfunction is linked with arterial stiffness across glucose metabolism: The Maastricht Study"

#### Removal of 24-hour HRV outliers

HRV indices are very sensitive to data error in the time-series collection of interbeat intervals. This can be due to missing heartbeat signals, arrhythmia, ectopic beats and more. These premature, skipped, or non-captured heartbeat recordings might not have been unfiltered from the IBI data and should have been removed, as it does not reflect autonomic activity. The time segment of frequency domain measures in the Maastricht Study was over the whole recording to capture very- and ultra-lower frequency components. Reference values of 24-hour HRV are done with small population size and with either time domain HRV or 5-min segments of frequency domain indices [1]. To determine cutoff for the exclusion of outliers, we visualize each HRV index value across ages and compare it to available reference values by age from studies [1]. Based on visually observing distribution and available reference material, we excluded time-domain indices according to reference material, and the upper 1$^{th}$ percentile in frequency-domain measures.

*1. Sammito S, Böckelmann I. Reference values for time- and frequency-domain heart rate variability measures. Heart Rhythm. 2016;13(6):1309-16. doi: 10.1016/j.hrthm.2016.02.006.*

#### Table 1S: Descriptives of included and non-included population

| **Characteristic** | **No**, N = 5,308 | **Yes**, N = 3,879 |
|---|---|---|
| Sex | | |
| Men | 2,782 (50%) | 1,789 (49%) |
| Women | 2,732 (50%) | 1,884 (51%) |
| Age (years) | 61 (53, 67) | 60 (53, 66) |
| Ethnicity | | |
| White | 5,418 (98%) | 3,633 (99%) |
| Non-white | 93 (1.7%) | 40 (1.1%) |
| Education | | |
| Low (No education, (un)completed primary education, or lower vocational education) | 1,911 (35%) | 1,094 (30%) |
| Middle (intermediate vocational education or higher secondary education) | 1,449 (27%) | 1,050 (29%) |
| High (Higher vocational education or university education) | 2,028 (38%) | 1,529 (42%) |
| Alcohol consumption | | |
| None | 1,083 (20%) | 609 (17%) |
| Low (Women: ≤ 7, Men: ≤ 14) | 3,237 (60%) | 2,147 (58%) |
| High (Women: > 7, Men: > 14) | 1,119 (21%) | 917 (25%) |
| Smoking status | | |
| Never | 2,086 (38%) | 1,417 (39%) |
| Former (quit > 6 months ago) | 2,511 (46%) | 1,733 (47%) |
| Former (quit < 6 months ago) | 106 (1.9%) | 62 (1.7%) |
| Current | 740 (14%) | 461 (13%) |
| Total physical activity (hours/week) | 13 (8, 18) | 13 (8, 19) |
| Moderate to vigorous physical activity (hours/week) | 4.5 (1.8, 7.5) | 4.5 (2.3, 7.8) |
| BMI ($kg^2/m$) | 26.4 (23.9, 29.5) | 26.0 (23.6, 28.8) |
| Waist (cm) | 95 (86, 105) | 93 (85, 102) |
| HbA1c (%) | 5.54 (5.26, 5.99) | 5.54 (5.26, 5.90) |
| Fasting plasma glucose (mmol/L) | 5.40 (5.00, 6.20) | 5.40 (4.90, 6.00) |
| LDL (mmol/L) | 2.90 (2.30, 3.70) | 3.10 (2.40, 3.80) |
| HDL (mmol/L) | 1.50 (1.20, 1.80) | 1.50 (1.20, 1.90) |
| Total cholesterol (mmol/L) | 5.10 (4.30, 5.90) | 5.30 (4.60, 6.10) |

| **Characteristic** | **No**, N = 5,308 | **Yes**, N = 3,879 |
|---|---|---|
| Triglycerides (mmol/L) | 1.19 (0.87, 1.70) | 1.18 (0.87, 1.65) |
| Glucose metabolism status | | |
| Normal glucose metabolism | 3,358 (61%) | 2,389 (65%) |
| Prediabetes | 844 (15%) | 538 (15%) |
| Type 2 Diabetes | 1,259 (23%) | 746 (20%) |
| Duration of type-2 diabetes (only for diagnosed participants) | 4 (0, 10) | 3 (0, 9) |
| Mean IBI (ms) | 829 (765, 910) | 828 (765, 904) |
| SDNN (ms) | 134 (110, 161) | 133 (110, 158) |
| RMSSD (ms) | 29 (21, 49) | 25 (20, 34) |
| SDANN (ms) | 121 (98, 148) | 119 (97, 143) |
| SDNNi (ms) | 54 (43, 71) | 52 (42, 63) |
| pNN50 (%) | 8 (3, 20) | 6 (3, 12) |
| TP ($ms^2$) | 11,509 (7,706, 16,540) | 11,566 (7,991, 16,394) |
| ULF ($ms^2$) | 9,665 (6,310, 14,055) | 9,788 (6,655, 14,183) |
| VLF ($ms^2$) | 1,064 (722, 1,606) | 1,105 (736, 1,571) |
| LF ($ms^2$) | 370 (213, 625) | 364 (222, 593) |
| HF ($ms^2$) | 104 (55, 212) | 84 (50, 149) |
| Systolic blood pressure (mmHg) | 126 (117, 136) | 126 (116, 136) |
| Diastolic blood pressure (mmHg) | 75 (71, 80) | 76 (71, 81) |
| Mean arterial pressure (mmHg) | 96 (89, 103) | 96 (89, 103) |
| Carotid artery distensibility (10-3/kPa) | 13.8 (10.7, 17.5) | 14.2 (11.0, 17.8) |
| Carotid-femoral pulse wave velocity (m/s) | 8.72 (7.60, 10.16) | 8.40 (7.44, 9.76) |
| Prior CVD | 1,482 (27%) | 0 (0%) |
| Hypertension (Yes) | 3,056 (56%) | 1,740 (47%) |
| Glucose lowering medication | 973 (18%) | 519 (14%) |
| Antihypertensive medication | 2,192 (40%) | 1,108 (30%) |

| **Characteristic** | **No**, N = 5,308 | **Yes**, N = 3,879 |
|---|---|---|
| Lipid-lowering medication | 1,803 (33%) | 905 (25%) |

n (%); Median (IQR)

#### Table 2S: Descriptives of participants with both CarDC and cf-PWV measured and participants with only cf-PWV measured

| Characteristic | **CAD measured**, N = 1,970 | **Without CAD measureMents**, N = 1,909 |
|---|---|---|
| Sex | | |
| Men | 901 (50%) | 888 (47%) |
| Women | 901 (50%) | 983 (53%) |
| Age (years) | 60 (54, 66) | 59 (52, 66) |
| Education | | |
| Low (No education, (un)completed primary education, or lower vocational education) | 506 (28%) | 588 (31%) |
| Middle (intermediate vocational education or higher secondary education) | 541 (30%) | 509 (27%) |
| High (Higher vocational education or university education) | 755 (42%) | 774 (41%) |
| Smoking status | | |
| Never | 643 (36%) | 774 (41%) |
| Former (quit > 6 months ago) | 892 (50%) | 841 (45%) |
| Former (quit < 6 months ago) | 33 (1.8%) | 29 (1.5%) |
| Current | 234 (13%) | 227 (12%) |
| BMI ($kg^2/m$) | 26.2 (23.7, 29.2) | 25.7 (23.5, 28.6) |
| Waist (cm) | 94 (86, 103) | 92 (83, 101) |
| HbA1c (%) | 5.63 (5.35, 5.99) | 5.44 (5.17, 5.72) |
| Fasting plasma glucose (mmol/L) | 5.50 (5.00, 6.30) | 5.30 (4.90, 5.80) |
| LDL (mmol/L) | 3.10 (2.40, 3.90) | 3.10 (2.40, 3.70) |
| HDL (mmol/L) | 1.50 (1.20, 1.80) | 1.60 (1.20, 1.90) |
| Total cholesterol (mmol/L) | 5.40 (4.60, 6.10) | 5.30 (4.60, 6.00) |
| Triglycerides (mmol/L) | 1.22 (0.89, 1.73) | 1.14 (0.84, 1.59) |
| Hypertension (Yes) | 928 (52%) | 812 (43%) |
| Diabetes status | | |
| Normal glucose metabolism | 1,049 (58%) | 1,340 (72%) |
| Prediabetes | 323 (18%) | 215 (11%) |
| Type 2 Diabetes | 430 (24%) | 316 (17%) |
| Duration of type-2 diabetes (only for diagnosed participants) | 3 (0, 8) | 3 (0, 9) |
| Mean IBI (ms) | 824 (759, 900) | 832 (772, 908) |

| **Characteristic** | **CAD measured**, N = 1,970 | **Without CAD measureMents**, N = 1,909 |
|---|---|---|
| SDNN (ms) | 132 (109, 157) | 133 (111, 159) |
| RMSSD (ms) | 25 (19, 34) | 26 (20, 34) |
| SDANN (ms) | 120 (96, 143) | 118 (98, 144) |
| SDNNi (ms) | 51 (42, 62) | 53 (43, 64) |
| pNN50 (%) | 6 (3, 12) | 6 (3, 12) |
| TP ($ms^2$) | 11,551 (7,860, 16,410) | 11,571 (8,088, 16,366) |
| ULF ($ms^2$) | 9,850 (6,503, 14,183) | 9,673 (6,780, 14,194) |
| VLF ($ms^2$) | 1,065 (707, 1,520) | 1,129 (767, 1,643) |
| LF ($ms^2$) | 350 (213, 578) | 381 (233, 613) |
| HF ($ms^2$) | 83 (48, 150) | 86 (52, 149) |
| Systolic blood pressure (mmHg) | 126 (117, 136) | 125 (115, 135) |
| Diastolic blood pressure (mmHg) | 76 (71, 81) | 75 (71, 81) |
| Mean arterial pressure (mmHg) | 96 (90, 103) | 96 (89, 103) |
| Carotid-femoral pulse wave velocity (m/s) | 8.48 (7.44, 9.84) | 8.32 (7.44, 9.60) |
| Glucose-lowering medication (Yes) | 301 (17%) | 218 (12%) |
| Antihypertensive medication (Yes) | 601 (33%) | 507 (27%) |
| Using beta-blockers (Yes) | 239 (13%) | 182 (9.7%) |

n (%); Median (IQR)

#### Table 3S: Study population characteristics by diabetes status

| Characteristic | **Normal glucose metabolism**, N = 2,546[1] | **Prediabetes**, N = 568[1] | **Type 2 Diabetes**, N = 765[1] |
|---|---|---|---|
| Sex | | | |
| Men | 1,028 (43%) | 280 (52%) | 481 (64%) |
| Women | 1,361 (57%) | 258 (48%) | 265 (36%) |
| Age (years) | 58 (51, 64) | 62 (57, 68) | 63 (57, 68) |
| Ethnicity | | | |
| White | 2,368 (99%) | 533 (99%) | 732 (98%) |
| Non-white | 21 (0.9%) | 5 (0.9%) | 14 (1.9%) |
| Education | | | |
| Low (No education, (un)completed primary education, or lower vocational education) | 604 (25%) | 192 (36%) | 298 (40%) |
| Middle (intermediate vocational education or higher secondary education) | 697 (29%) | 145 (27%) | 208 (28%) |
| High (Higher vocational education or university education) | 1,088 (46%) | 201 (37%) | 240 (32%) |
| Alcohol consumption | | | |
| None | 338 (14%) | 83 (15%) | 188 (25%) |
| Low (Women: ≤ 7, Men: ≤ 14) | 1,437 (60%) | 298 (55%) | 412 (55%) |
| High (Women: > 7, Men: > 14) | 614 (26%) | 157 (29%) | 146 (20%) |
| Smoking status | | | |
| Never | 988 (41%) | 185 (34%) | 244 (33%) |
| Former (quit > 6 months ago) | 1,070 (45%) | 286 (53%) | 377 (51%) |
| Former (quit < 6 months ago) | 43 (1.8%) | 3 (0.6%) | 16 (2.1%) |
| Current | 288 (12%) | 64 (12%) | 109 (15%) |
| Total physical activity (hours/week) | 13 (9, 19) | 13 (9, 19) | 12 (7, 17) |
| Moderate to vigorous physical activity (hours/week) | 5.3 (3.0, 8.3) | 4.5 (2.3, 7.5) | 3.8 (1.5, 6.8) |
| BMI (kg²/m) | 25.0 (22.9, 27.4) | 27.2 (24.9, 30.1) | 28.8 (26.0, 31.7) |

| **Characteristic** | **Normal glucose metabolism**, N = 2,546[1] | **Prediabetes**, N = 568[1] | **Type 2 Diabetes**, N = 765[1] |
|---|---|---|---|
| Waist (cm) | 89 (81, 97) | 98 (90, 105) | 103 (96, 112) |
| HbA1c (%) | 5.35 (5.17, 5.63) | 5.63 (5.35, 5.90) | 6.54 (6.08, 7.09) |
| Fasting plasma glucose (mmol/L) | 5.10 (4.80, 5.40) | 5.90 (5.40, 6.30) | 7.40 (6.60, 8.50) |
| LDL (mmol/L) | 3.20 (2.70, 3.90) | 3.30 (2.60, 4.00) | 2.40 (1.80, 3.10) |
| HDL (mmol/L) | 1.60 (1.30, 2.00) | 1.40 (1.20, 1.80) | 1.30 (1.00, 1.50) |
| Total cholesterol (mmol/L) | 5.50 (4.80, 6.20) | 5.50 (4.80, 6.30) | 4.50 (3.90, 5.20) |
| Triglycerides (mmol/L) | 1.05 (0.80, 1.45) | 1.39 (1.03, 1.90) | 1.51 (1.08, 2.14) |
| Duration of type-2 diabetes (only for diagnosed participants) | NA (NA, NA) | NA (NA, NA) | 3 (0, 9) |
| Mean IBI (ms) | 838 (775, 907) | 815 (760, 897) | 806 (744, 889) |
| SDNN (ms) | 138 (117, 164) | 127 (106, 152) | 116 (96, 139) |
| RMSSD (ms) | 26 (21, 34) | 24 (19, 33) | 22 (17, 31) |
| SDANN (ms) | 125 (103, 149) | 113 (92, 139) | 103 (84, 127) |
| SDNNi (ms) | 55 (46, 65) | 50 (41, 60) | 44 (36, 54) |
| pNN50 (%) | 7 (3, 13) | 5 (2, 10) | 4 (2, 9) |
| TP ($ms^2$) | 12,596 (8,880, 17,498) | 10,615 (7,134, 15,374) | 8,880 (6,064, 12,722) |
| ULF ($ms^2$) | 10,771 (7,392, 15,142) | 8,948 (5,852, 13,374) | 7,524 (5,036, 11,001) |
| VLF ($ms^2$) | 1,198 (833, 1,692) | 1,015 (685, 1,478) | 816 (541, 1,267) |
| LF ($ms^2$) | 421 (257, 651) | 328 (200, 540) | 261 (154, 422) |
| HF ($ms^2$) | 94 (57, 158) | 78 (47, 138) | 63 (36, 117) |
| Systolic blood pressure (mmHg) | 123 (114, 133) | 129 (122, 140) | 130 (122, 139) |
| Diastolic blood pressure (mmHg) | 75 (71, 80) | 78 (73, 83) | 76 (72, 81) |
| Mean arterial pressure (mmHg) | 95 (88, 102) | 99 (93, 107) | 98 (92, 105) |
| Carotid artery distensibility (10-3/kPa) | 15.0 (11.8, 18.8) | 13.5 (10.4, 16.9) | 12.5 (9.9, 16.0) |
| Carotid-femoral pulse wave velocity (m/s) | 8.08 (7.28, 9.16) | 8.96 (7.84, 10.32) | 9.36 (8.16, 10.80) |

| **Characteristic** | **Normal glucose metabolism**, N = 2,546[1] | **Prediabetes**, N = 568[1] | **Type 2 Diabetes**, N = 765[1] |
|---|---|---|---|
| Hypertension (Yes) | 833 (35%) | 317 (59%) | 590 (79%) |
| Glucose-lowering medication (Yes) | 0 (0%) | 0 (0%) | 519 (70%) |
| Antihypertensive medication (Yes) | 431 (18%) | 199 (37%) | 478 (64%) |
| Using beta-blockers (Yes) | 149 (6.2%) | 77 (14%) | 195 (26%) |
| Lipid-lowering medication | 280 (12%) | 141 (26%) | 484 (65%) |

[1]n (%); Median (IQR)

#### Table 4S: Association between long-term HRV in the original unit and pulse wave velocity

| | *Model 1* | *Model 2* |
|---|---|---|
| **HRV index** | **PWV % (95% CI)** | **PWV % (95% CI)** |
| Mean IBI (ms) | -0.02255 (-0.02769; -0.017) | -0.02249 (-0.02771; -0.017) |
| SDNN (ms) | -0.07141 (-0.08689; -0.056) | -0.07189 (-0.08769; -0.056) |
| SDANN (ms) | -0.06527 (-0.08119; -0.049) | -0.06548 (-0.08166; -0.049) |
| SDNNi (ms) | -0.16548 (-0.20319; -0.128) | -0.16403 (-0.20266; -0.125) |
| RMSSD (ms) | -0.09565 (-0.13802; -0.053) | -0.09239 (-0.13495; -0.05) |
| pNN50 (%) | -0.13984 (-0.20466; -0.075) | -0.13592 (-0.20105; -0.071) |
| TP (ms²) | -0.00035 (-0.00043; 0.000) | -0.00035 (-0.00043; 0000) |
| HF (ms²) | -0.00857 (-0.01357; -0.004) | -0.00826 (-0.01326; -0.003) |
| LF (ms²) | -0.00445 (-0.00627; -0.003) | -0.00427 (-0.00612; -0.002) |
| VLF (ms²) | -0.0034 (-0.00418; -0.003) | -0.00337 (-0.00416; -0.003) |
| ULF (ms²) | -0.00035 (-0.00044; 0000) | -0.00035 (-0.00045; 0000) |

*Percentage CD per original unit increase in heart rate variability index and heart period intervals. Model 1: adjusted for sex, age, educational status, diabetes status, and mean arterial pressure. Model 2: Model 1 + physical activity, smoking behaviour, alcohol use, body mass index, hba1c, triglycerides, total-to-high density lipoprotein cholesterol ratio, lipid-modifying- and antihypertensive medication.*

#### Table 5S: Association between long-term HRV in the original unit and carotid distensibility

| | Model 1 | Model 2 |
|---|---|---|
| **HRV index** | **CD % (95% CI)** | **CD % (95% CI)** |
| Mean IBI (ms) | 0.0409 (0.02745; 0.054) | 0.04214 (0.02858; 0.056) |
| SDNN (ms) | 0.08919 (0.04849; 0.130) | 0.08917 (0.04786; 0.130) |
| SDANN (ms) | 0.08196 (0.04018; 0.124) | 0.0814 (0.0391; 0.124) |
| SDNNi (ms) | 0.18749 (0.08769; 0.287) | 0.19231 (0.09109; 0.294) |
| RMSSD (ms) | 0.07784 (-0.02725; 0.183) | 0.09135 (-0.01362; 0.196) |
| pNN50 | 0.10975 (-0.05103; 0.271) | 0.12704 (-0.03355; 0.288) |
| TP ($ms^2$) | 0.00041 (0.00019; 0.001) | 0.00041 (0.00019; 0.001) |
| HF ($ms^2$) | 0.00415 (-0.00829; 0.017) | 0.00523 (-0.00716; 0.018) |
| LF ($ms^2$) | 0.00575 (0.00088; 0.011) | 0.00572 (0.00082; 0.011) |
| VLF ($ms^2$) | 0.00327 (0.00118; 0.005) | 0.00336 (0.00124; 0.005) |
| ULF ($ms^2$) | 0.00043 (0.00019; 0.001) | 0.00043 (0.00019; 0.001) |

*Percentage CD per original unit increase in heart rate variability index and heart period intervals. Model 1: adjusted for sex, age, educational status, diabetes status, and mean arterial pressure. Model 2: Model 1 + physical activity, smoking behaviour, alcohol use, body mass index, hba1c, triglycerides, total-to-high density lipoprotein cholesterol ratio, lipid-modifying- and antihypertensive medication.*

### Table 6S: Association between long-term standardized HRV and pulse wave velocity by diabetes status

| | PWV % (95% CI) | Interaction p-value |
|---|---|---|
| **Heart period (ms)** | | |
| Normal glucose metabolism | -2.542 (-3.213; -1.866) | ref. |
| Prediabetes | -1.85 (-3.163; -0.519) | 0.35031 |
| Type 2 Diabetes | -2.225 (-3.313; -1.126) | 0.62224 |
| **SDNN (ms)** | | |
| Normal glucose metabolism | -1.745 (-2.406; -1.08) | ref. |
| Prediabetes | -4.397 (-5.763; -3.012) | 0.00065 |
| Type 2 Diabetes | -3.725 (-4.899; -2.537) | 0.00402 |
| **SDANN (ms)** | | |
| Normal glucose metabolism | -1.491 (-2.147; -0.831) | ref. |
| Prediabetes | -4.012 (-5.361; -2.643) | 0.00108 |
| Type 2 Diabetes | -3.291 (-4.47; -2.097) | 0.00916 |
| **SDNN index (ms)** | | |
| Normal glucose metabolism | -1.851 (-2.552; -1.146) | ref. |
| Prediabetes | -3.914 (-5.264; -2.544) | 0.00692 |
| Type 2 Diabetes | -3.272 (-4.43; -2.1) | 0.03746 |
| **RMSSD (ms)** | | |
| Normal glucose metabolism | -0.967 (-1.651; -0.278) | ref. |
| Prediabetes | -2.011 (-3.269; -0.738) | 0.15392 |
| Type 2 Diabetes | -0.995 (-2.042; 0.063) | 0.96501 |
| **pNN50 (%)** | | |
| Normal glucose metabolism | -0.974 (-1.647; -0.297) | ref. |
| Prediabetes | -1.68 (-2.944; -0.4) | 0.33419 |
| Type 2 Diabetes | -1.095 (-2.241; 0.063) | 0.85863 |

| | PWV % (95% CI) | Interaction p-value |
|---|---|---|
| **Time-domain Z-score** | | |
| Normal glucose metabolism | -2.053 (-2.863; -1.236) | ref. |
| Prediabetes | -4.897 (-6.523; -3.243) | 0.00222 |
| Type 2 Diabetes | -3.467 (-4.83; -2.084) | 0.08088 |
| **Total power (ms²)** | | |
| Normal glucose metabolism | -1.57 (-2.206; -0.93) | ref. |
| Prediabetes | -4.336 (-5.735; -2.916) | 0.00046 |
| Type 2 Diabetes | -3.451 (-4.753; -2.132) | 0.01121 |
| **HF (ms²)** | | |
| Normal glucose metabolism | -0.538 (-1.205; 0.133) | ref. |
| Prediabetes | -1.882 (-3.234; -0.511) | 0.08129 |
| Type 2 Diabetes | -1.249 (-2.382; -0.103) | 0.29055 |
| **LF (ms²)** | | |
| Normal glucose metabolism | -0.7 (-1.391; -0.004) | ref. |
| Prediabetes | -3.425 (-4.795; -2.035) | 4e-04 |
| Type 2 Diabetes | -2.144 (-3.415; -0.855) | 0.04795 |
| **VLF (ms²)** | | |
| Normal glucose metabolism | -1.946 (-2.605; -1.282) | ref. |
| Prediabetes | -3.736 (-5.092; -2.36) | 0.01869 |
| Type 2 Diabetes | -3.243 (-4.537; -1.931) | 0.07796 |
| **ULF (ms²)** | | |
| Normal glucose metabolism | -1.442 (-2.072; -0.807) | ref. |
| Prediabetes | -3.975 (-5.364; -2.566) | 0.00125 |
| Type 2 Diabetes | -3.273 (-4.582; -1.946) | 0.01403 |
| **Frequency-domain Z-score** | | |

| | PWV % (95% CI) | Interaction p-value |
|---|---|---|
| Normal glucose metabolism | -1.968 (-2.778; -1.153) | ref. |
| Prediabetes | -5.652 (-7.367; -3.904) | 0.00015 |
| Type 2 Diabetes | -3.871 (-5.386; -2.333) | 0.02979 |

*Percentage PWV per SD increase in heart rate variability index and heart period intervals Adjustment from model 2 including age, sex, education, mean arterial pressure, physical activity, smoking, alcohol, body mass index, triglycerides, total-to-high density lipoprotein cholesterol ratio, lipid-modifying- and antihypertensive medication.*

**Table 7S: Association between long-term standardized HRV and carotid distensibility by diabetes status**

| | CD % (95% CI) | Interaction p-value |
|---|---|---|
| **Heart period (ms)** | | |
| Normal glucose metabolism | 3.755 (1.818; 5.729) | ref. |
| Prediabetes | 5.513 (2.119; 9.021) | 0.37357 |
| Type 2 Diabetes | 5.663 (2.848; 8.555) | 0.26882 |
| **SDNN (ms)** | | |
| Normal glucose metabolism | 1.223 (-0.643; 3.124) | ref. |
| Prediabetes | 7.175 (3.577; 10.898) | 0.00349 |
| Type 2 Diabetes | 5.358 (2.347; 8.458) | 0.0214 |
| **SDANN (ms)** | | |
| Normal glucose metabolism | 1.16 (-0.691; 3.044) | ref. |
| Prediabetes | 5.728 (2.293; 9.277) | 0.02074 |
| Type 2 Diabetes | 4.76 (1.749; 7.86) | 0.04525 |
| **SDNN index (ms)** | | |
| Normal glucose metabolism | 1.338 (-0.632; 3.348) | ref. |
| Prediabetes | 6.916 (3.28; 10.681) | 0.00651 |
| Type 2 Diabetes | 3.944 (0.981; 6.993) | 0.14548 |
| **RMSSD (ms)** | | |
| Normal glucose metabolism | 0.311 (-1.461; 2.115) | ref. |
| Prediabetes | 3.05 (0.15; 6.034) | 0.11407 |
| Type 2 Diabetes | 1.386 (-1.187; 4.026) | 0.50357 |
| **pNN50 (%)** | | |
| Normal glucose metabolism | 0.15 (-1.589; 1.919) | ref. |
| Prediabetes | 2.933 (0.015; 5.936) | 0.10757 |
| Type 2 Diabetes | 1.64 (-1.131; 4.489) | 0.37422 |

| | | |
|---|---|---|
| **Time-domain Z-score** | | |
| Normal glucose metabolism | 1.223 (-1.033; 3.53) | ref. |
| Prediabetes | 8.049 (3.778; 12.495) | 0.00486 |
| Type 2 Diabetes | 4.808 (1.316; 8.421) | 0.09046 |
| **Total power (ms²)** | | |
| Normal glucose metabolism | 1.087 (-0.708; 2.915) | ref. |
| Prediabetes | 6.053 (2.429; 9.807) | 0.015 |
| Type 2 Diabetes | 5.35 (2.026; 8.782) | 0.0261 |
| **HF (ms²)** | | |
| Normal glucose metabolism | -0.517 (-2.228; 1.223) | ref. |
| Prediabetes | 3.06 (-0.032; 6.247) | 0.04629 |
| Type 2 Diabetes | 1.397 (-1.375; 4.247) | 0.25123 |
| **LF (ms²)** | | |
| Normal glucose metabolism | 0.892 (-1.027; 2.849) | ref. |
| Prediabetes | 4.128 (0.516; 7.869) | 0.11249 |
| Type 2 Diabetes | 2.689 (-0.613; 6.101) | 0.34982 |
| **VLF (ms²)** | | |
| Normal glucose metabolism | 1.258 (-0.641; 3.193) | ref. |
| Prediabetes | 5.66 (2.01; 9.442) | 0.03227 |
| Type 2 Diabetes | 3.494 (0.206; 6.89) | 0.24368 |
| **ULF (ms²)** | | |
| Normal glucose metabolism | 0.996 (-0.778; 2.801) | ref. |
| Prediabetes | 5.535 (1.98; 9.213) | 0.0241 |
| Type 2 Diabetes | 5.373 (2.027; 8.828) | 0.02287 |
| **Frequency-domain Z-score** | | |
| Normal glucose metabolism | 1.186 (-1.098; 3.523) | ref. |
| Prediabetes | 8.277 (3.61; 13.154) | 0.0063 |

| | | |
|---|---|---|
| Type 2 Diabetes | 5.313 (1.374; 9.405) | 0.07353 |

*Percentage CD per SD increase in heart rate variability index and heart period intervals Adjustment from model 2 including age, sex, education, mean arterial pressure, physical activity, smoking, alcohol, body mass index, triglycerides, total-to-high density lipoprotein cholesterol ratio, lipid-modifying- and antihypertensive medication.*

#### Table 8S: Association between long-term standardized HRV and pulse wave velocity by sex

| | PWV % (95% CI) | Interaction p-value |
|---|---|---|
| **Heart period (ms)** | | |
| Men | -1.853 (-2.538; -1.162) | ref. |
| Women | -3.159 (-3.997; -2.314) | 0.01635 |
| **SDNN (ms)** | | |
| Men | -2.377 (-3.109; -1.639) | ref. |
| Women | -2.646 (-3.398; -1.889) | 0.60266 |
| **SDANN (ms)** | | |
| Men | -2.095 (-2.829; -1.355) | ref. |
| Women | -2.312 (-3.055; -1.562) | 0.67666 |
| **SDNN index (ms)** | | |
| Men | -2.041 (-2.769; -1.306) | ref. |
| Women | -3.012 (-3.823; -2.194) | 0.06509 |
| **RMSSD (ms)** | | |
| Men | -1.012 (-1.711; -0.308) | ref. |
| Women | -1.316 (-2.093; -0.534) | 0.56598 |
| **pNN50 (%)** | | |
| Men | -1.059 (-1.797; -0.316) | ref. |
| Women | -1.18 (-1.924; -0.43) | 0.81918 |
| **Time-domain Z-score** | | |
| Men | -2.501 (-3.372; -1.622) | ref. |
| Women | -3.062 (-3.98; -2.134) | 0.37062 |
| **Total power (ms²)** | | |
| Men | -2.044 (-2.777; -1.305) | ref. |
| Women | -2.432 (-3.18; -1.678) | 0.45651 |
| **HF (ms²)** | | |
| Men | -0.717 (-1.476; 0.048) | ref. |
| Women | -1.049 (-1.782; -0.312) | 0.532 |
| **LF (ms²)** | | |
| Men | -0.941 (-1.637; -0.24) | ref. |
| Women | -2.126 (-3.034; -1.209) | 0.03287 |
| **VLF (ms²)** | | |
| Men | -2.04 (-2.729; -1.345) | ref. |
| Women | -3.027 (-3.884; -2.164) | 0.06757 |
| **ULF (ms²)** | | |
| Men | -1.903 (-2.639; -1.162) | ref. |
| Women | -2.232 (-2.971; -1.486) | 0.52879 |
| **Frequency-domain Z-score** | | |
| Men | -2.407 (-3.293; -1.512) | ref. |
| Women | -3.266 (-4.226; -2.297) | 0.17941 |

*Percentage PWV per SD increase in heart rate variability index and heart period intervals. Adjustment from model 2 including age, education, diabetes status, mean arterial pressure, physical activity, smoking, alcohol, body mass index, triglycerides, total-to-high density lipoprotein cholesterol ratio, lipid-modifying- and antihypertensive medication.*

#### Table 9S: Association between long-term standardized HRV and carotid distensibility by sex

| | CD % (95% CI) | Interaction p-value |
|---|---|---|
| **Heart period (ms)** | | |
| Men | 5.87 (3.977; 7.797) | ref. |
| Women | 2.489 (0.177; 4.853) | 0.02581 |
| **SDNN (ms)** | | |
| Men | 4.006 (2.021; 6.03) | ref. |
| Women | 2.23 (0.136; 4.368) | 0.21613 |
| **SDANN (ms)** | | |
| Men | 3.228 (1.262; 5.232) | ref. |
| Women | 2.293 (0.227; 4.402) | 0.51292 |
| **SDNN index (ms)** | | |
| Men | 4.253 (2.26; 6.285) | ref. |
| Women | 1.113 (-1.13; 3.407) | 0.03247 |
| **RMSSD (ms)** | | |
| Men | 2.788 (1.024; 4.582) | ref. |
| Women | -0.922 (-2.846; 1.04) | 0.00546 |
| **pNN50 (%)** | | |
| Men | 2.707 (0.848; 4.6) | ref. |
| Women | -0.65 (-2.496; 1.232) | 0.01171 |
| **Time-domain Z-score** | | |
| Men | 5.021 (2.662; 7.436) | ref. |
| Women | 1.141 (-1.365; 3.711) | 0.02393 |
| **Total power ($ms^2$)** | | |
| Men | 3.186 (1.209; 5.2) | ref. |
| Women | 2.147 (0.045; 4.292) | 0.47122 |
| **HF ($ms^2$)** | | |
| Men | 2.407 (0.523; 4.325) | ref. |
| Women | -1.254 (-3.089; 0.616) | 0.00618 |
| **LF ($ms^2$)** | | |
| Men | 2.606 (0.698; 4.55) | ref. |
| Women | 0.298 (-2.171; 2.83) | 0.13317 |
| **VLF ($ms^2$)** | | |
| Men | 3.274 (1.366; 5.218) | ref. |
| Women | 0.974 (-1.466; 3.473) | 0.13568 |
| **ULF ($ms^2$)** | | |
| Men | 2.881 (0.907; 4.893) | ref. |

| | CD % (95% CI) | Interaction p-value |
|---|---|---|
| Women | 2.198 (0.134; 4.305) | 0.6343 |
| **Frequency-domain Z-score** | | |
| Men | 4.491 (2.058; 6.981) | ref. |
| Women | 1.389 (-1.31; 4.162) | 0.0849 |

Percentage CD per SD increase in heart rate variability index and heart period intervals. *Adjustment from model 2 including age, education, diabetes status, mean arterial pressure, physical activity, smoking, alcohol, body mass index, triglycerides, total-to-high density lipoprotein cholesterol ratio, lipid-modifying- and antihypertensive medication.*

#### Table 10S: Sensitivity analysis: Association between long-term HRV and pulse wave velocity

| Sub-group | Population size | PWV % | 5% | 95% |
|---|---|---|---|---|
| **Heart period (ms)** | | | | |
| Main | 3673 | -0.0225 | -0.0277 | -0.0173 |
| No beta-blocker medication | 3252 | -0.0208 | -0.0265 | -0.0151 |
| No antihypertension medication | 2565 | -0.0189 | -0.0250 | -0.0127 |
| No diabetes and antihypertension medication | 1958 | -0.0199 | -0.0266 | -0.0133 |
| **SDNN (ms)** | | | | |
| Main | 3673 | -0.0719 | -0.0877 | -0.0561 |
| No beta-blocker medication | 3252 | -0.0701 | -0.0864 | -0.0537 |
| No antihypertension medication | 2565 | -0.0642 | -0.0820 | -0.0465 |
| No diabetes and antihypertension medication | 1958 | -0.0509 | -0.0700 | -0.0319 |
| **SDANN (ms)** | | | | |
| Main | 3673 | -0.0655 | -0.0816 | -0.0493 |
| No beta-blocker medication | 3252 | -0.0641 | -0.0808 | -0.0474 |
| No antihypertension medication | 2565 | -0.0586 | -0.0767 | -0.0404 |
| No diabetes and antihypertension medication | 1958 | -0.0443 | -0.0638 | -0.0248 |
| **SDNN index (ms)** | | | | |
| Main | 3673 | -0.1639 | -0.2026 | -0.1253 |

| Sub-group | Population size | PWV % | 5% | 95% |
|---|---|---|---|---|
| No beta-blocker medication | 3252 | -0.1657 | -0.2063 | -0.1250 |
| No antihypertension medication | 2565 | -0.1476 | -0.1914 | -0.1038 |
| No diabetes and antihypertension medication | 1958 | -0.1311 | -0.1786 | -0.0836 |
| **RMSSD (ms)** | | | | |
| Main | 3673 | -0.0923 | -0.1349 | -0.0498 |
| No beta-blocker medication | 3252 | -0.1111 | -0.1564 | -0.0657 |
| No antihypertension medication | 2565 | -0.1045 | -0.1546 | -0.0542 |
| No diabetes and antihypertension medication | 1958 | -0.0932 | -0.1486 | -0.0377 |
| **pNN50 (%)** | | | | |
| Main | 3673 | -0.1359 | -0.2010 | -0.0707 |
| No beta-blocker medication | 3252 | -0.1546 | -0.2233 | -0.0859 |
| No antihypertension medication | 2565 | -0.1520 | -0.2264 | -0.0776 |
| No diabetes and antihypertension medication | 1958 | -0.1510 | -0.2330 | -0.0689 |
| **Total power ($ms^2$)** | | | | |
| Main | 3673 | -0.0003 | -0.0004 | -0.0003 |
| No beta-blocker medication | 3252 | -0.0003 | -0.0004 | -0.0003 |
| No antihypertension medication | 2565 | -0.0003 | -0.0004 | -0.0002 |
| No diabetes and antihypertension medication | 1958 | -0.0002 | -0.0003 | -0.0002 |
| **HF ($ms^2$)** | | | | |

| Sub-group | Population size | PWV % | 5% | 95% |
|---|---|---|---|---|
| Main | 3673 | -0.0083 | -0.0133 | -0.0033 |
| No beta-blocker medication | 3252 | -0.0096 | -0.0148 | -0.0043 |
| No antihypertension medication | 2565 | -0.0088 | -0.0145 | -0.0032 |
| No diabetes and antihypertension medication | 1958 | -0.0066 | -0.0128 | -0.0004 |
| **LF ($ms^2$)** | | | | |
| Main | 3673 | -0.0043 | -0.0061 | -0.0024 |
| No beta-blocker medication | 3252 | -0.0046 | -0.0065 | -0.0026 |
| No antihypertension medication | 2565 | -0.0040 | -0.0061 | -0.0020 |
| No diabetes and antihypertension medication | 1958 | -0.0033 | -0.0054 | -0.0011 |
| **VLF ($ms^2$)** | | | | |
| Main | 3673 | -0.0034 | -0.0042 | -0.0026 |
| No beta-blocker medication | 3252 | -0.0033 | -0.0042 | -0.0025 |
| No antihypertension medication | 2565 | -0.0031 | -0.0039 | -0.0022 |
| No diabetes and antihypertension medication | 1958 | -0.0029 | -0.0038 | -0.0020 |
| **ULF ($ms^2$)** | | | | |
| Main | 3673 | -0.0004 | -0.0004 | -0.0003 |
| No beta-blocker medication | 3252 | -0.0003 | -0.0004 | -0.0003 |
| No antihypertension medication | 2565 | -0.0003 | -0.0004 | -0.0002 |
| No diabetes and antihypertension medication | 1958 | -0.0002 | -0.0004 | -0.0001 |

Percentage PWV per original unit increase in heart rate variability index and heart period intervals

Main: Model 2 (adjusted for antihypertensive medication)

No antihypertensive medication: people with antihypertensive medication was excluded

No antihypertensive medication and without diabetes: people with antihypertensive medication and diabetes was excluded

#### Table 11S: Sensitivity analysis: Association between long-term HRV (in original unit) and carotid distensibility

| Sub-group | Population size | CD % | 5% | 95% |
|---|---|---|---|---|
| **Heart period (ms)** | | | | |
| Main | 1802 | 0.0420 | 0.0284 | 0.0555 |
| No beta-blocker medication | 1563 | 0.0402 | 0.0252 | 0.0553 |
| No antihypertension medication | 1201 | 0.0366 | 0.0201 | 0.0530 |
| No diabetes and antihypertension medication | 846 | 0.0289 | 0.0092 | 0.0485 |
| **SDNN (ms)** | | | | |
| Main | 1802 | 0.0887 | 0.0474 | 0.1300 |
| No beta-blocker medication | 1563 | 0.0968 | 0.0529 | 0.1407 |
| No antihypertension medication | 1201 | 0.0874 | 0.0388 | 0.1361 |
| No diabetes and antihypertension medication | 846 | 0.0499 | -0.0077 | 0.1076 |
| **SDANN (ms)** | | | | |
| Main | 1802 | 0.0809 | 0.0387 | 0.1232 |
| No beta-blocker medication | 1563 | 0.0913 | 0.0465 | 0.1361 |
| No antihypertension medication | 1201 | 0.0852 | 0.0353 | 0.1352 |
| No diabetes and antihypertension medication | 846 | 0.0572 | -0.0017 | 0.1161 |
| **SDNN index (ms)** | | | | |
| Main | 1802 | 0.1902 | 0.0890 | 0.2915 |
| No beta-blocker medication | 1563 | 0.1772 | 0.0670 | 0.2876 |
| No antihypertension medication | 1201 | 0.1515 | 0.0317 | 0.2714 |
| No diabetes and antihypertension medication | 846 | 0.0695 | -0.0728 | 0.2120 |
| **RMSSD (ms)** | | | | |
| Main | 1802 | 0.0915 | -0.0134 | 0.1965 |
| No beta-blocker medication | 1563 | 0.0712 | -0.0444 | 0.1869 |
| No antihypertension medication | 1201 | 0.1012 | -0.0289 | 0.2315 |
| No diabetes and antihypertension medication | 846 | 0.0436 | -0.1123 | 0.1998 |
| **pNN50 (%)** | | | | |
| Main | 1802 | 0.1274 | -0.0330 | 0.2881 |
| No beta-blocker medication | 1563 | 0.1098 | -0.0630 | 0.2829 |
| No antihypertension medication | 1201 | 0.1362 | -0.0555 | 0.3283 |
| No diabetes and antihypertension medication | 846 | 0.0560 | -0.1743 | 0.2868 |
| **Total power ($ms^2$)** | | | | |
| Main | 1802 | 0.0004 | 0.0002 | 0.0006 |
| No beta-blocker medication | 1563 | 0.0004 | 0.0002 | 0.0007 |
| No antihypertension medication | 1201 | 0.0004 | 0.0002 | 0.0007 |
| No diabetes and antihypertension medication | 846 | 0.0003 | 0.0000 | 0.0006 |
| **HF ($ms^2$)** | | | | |
| Main | 1802 | 0.0053 | -0.0070 | 0.0177 |

| | | | | |
|---|---|---|---|---|
| No beta-blocker medication | 1563 | 0.0045 | -0.0087 | 0.0177 |
| No antihypertension medication | 1201 | 0.0099 | -0.0047 | 0.0246 |
| No diabetes and antihypertension medication | 846 | 0.0038 | -0.0138 | 0.0214 |
| **LF (ms²)** | | | | |
| Main | 1802 | 0.0056 | 0.0007 | 0.0105 |
| No beta-blocker medication | 1563 | 0.0047 | -0.0006 | 0.0100 |
| No antihypertension medication | 1201 | 0.0043 | -0.0013 | 0.0099 |
| No diabetes and antihypertension medication | 846 | 0.0017 | -0.0048 | 0.0081 |
| **VLF (ms²)** | | | | |
| Main | 1802 | 0.0033 | 0.0012 | 0.0054 |
| No beta-blocker medication | 1563 | 0.0031 | 0.0008 | 0.0054 |
| No antihypertension medication | 1201 | 0.0026 | 0.0002 | 0.0050 |
| No diabetes and antihypertension medication | 846 | 0.0010 | -0.0018 | 0.0038 |
| **ULF (ms²)** | | | | |
| Main | 1802 | 0.0004 | 0.0002 | 0.0007 |
| No beta-blocker medication | 1563 | 0.0005 | 0.0002 | 0.0007 |
| No antihypertension medication | 1201 | 0.0004 | 0.0002 | 0.0007 |
| No diabetes and antihypertension medication | 846 | 0.0003 | 0.0000 | 0.0006 |

Percentage CD per original unit increase in heart rate variability index and mean heart period intervals

Main: model 2 (adjusted for antihypertensive medication)

No antihypertensive medication: people with antihypertensive medication was excluded

No antihypertensive medication and without diabetes: people with antihypertensive medication and diabetes was excluded

#### Table 12S: Association between long-term standardized HRV and pulse wave velocity

| | Model 1 | Model 2 |
|---|---|---|
| HRV index | PWV % (95% CI) | PWV % (95% CI) |
| Mean IBI (ms) | -2.373 (-2.906; -1.838) | -2.366 (-2.908; -1.822) |
| SDNN (ms) | -2.492 (-3.024; -1.957) | -2.508 (-3.051; -1.962) |
| SDANN (ms) | -2.195 (-2.724; -1.664) | -2.202 (-2.739; -1.662) |
| SDNNi (ms) | -2.487 (-3.045; -1.925) | -2.465 (-3.037; -1.890) |
| RMSSD (ms) | -1.189 (-1.711; -0.664) | -1.148 (-1.673; -0.621) |
| pNN50 | -1.151 (-1.681; -0.619) | -1.119 (-1.652; -0.584) |
| Time-domain Z-score | -2.787 (-3.431; -2.139) | -2.766 (-3.42; -2.106) |
| TP (ms²) | -2.235 (-2.766; -1.701) | -2.234 (-2.773; -1.692) |
| HF (ms²) | -0.922 (-1.456; -0.385) | -0.888 (-1.423; -0.35) |
| LF (ms²) | -1.412 (-1.984; -0.836) | -1.357 (-1.938; -0.773) |
| VLF (ms²) | -2.442 (-2.990; -1.890) | -2.416 (-2.975; -1.854) |
| ULF (ms²) | -2.069 (-2.596; -1.539) | -2.067 (-2.601; -1.529) |
| Frequency-domain Z-score | -2.819 (-3.487; -2.146) | -2.798 (-3.477; -2.113) |

*Percentage PWV per SD increase in heart rate variability index and heart period intervals Model 1: adjusted for sex, age, educational status, diabetes status, and mean arterial pressure. Model 2: Model 1 + physical activity, smoking behaviour, alcohol use, body mass index, hba1c, triglycerides, total-to-high density lipoprotein cholesterol ratio, lipid-modifying- and antihypertensive medication.*

.

#### Table 13S: Association between long-term standardized HRV and carotid distensibility

| | Model 1 | Model 2 |
|---|---|---|
| HRV index | CD % (95% CI) | CD % (95% CI) |
| Mean IBI (ms) | 4.45 (2.966; 5.956) | 4.588 (3.09; 6.107) |
| SDNN (ms) | 3.199 (1.727; 4.692) | 3.198 (1.705; 4.714) |
| SDANN (ms) | 2.824 (1.375; 4.294) | 2.805 (1.338; 4.293) |
| SDNNi (ms) | 2.889 (1.342; 4.46) | 2.964 (1.394; 4.559) |
| RMSSD (ms) | 0.977 (-0.34; 2.312) | 1.148 (-0.17; 2.483) |
| pNN50 | 0.912 (-0.422; 2.263) | 1.056 (-0.277; 2.407) |
| Time-domain Z-score | 3.162 (1.397; 4.959) | 3.284 (1.500; 5.100) |
| TP ($ms^2$) | 2.696 (1.226; 4.187) | 2.724 (1.237; 4.232) |
| HF ($ms^2$) | 0.449 (-0.892; 1.809) | 0.567 (-0.77; 1.922) |
| LF ($ms^2$) | 1.857 (0.282; 3.457) | 1.847 (0.263; 3.455) |
| VLF ($ms^2$) | 2.405 (0.858; 3.976) | 2.471 (0.908; 4.058) |
| ULF ($ms^2$) | 2.564 (1.112; 4.037) | 2.574 (1.108; 4.061) |
| Frequency-domain Z-score | 3.098 (1.229; 5.001) | 3.184 (1.295; 5.109) |

*Percentage CD per SD increase in heart rate variability index and heart period intervals. Model 1: adjusted for sex, age, educational status, diabetes status, and mean arterial pressure. Model 2: Model 1 + physical activity, smoking behaviour, alcohol use, body mass index, hba1c, triglycerides, total-to-high density lipoprotein cholesterol ratio, lipid-modifying- and antihypertensive medication.*

#### Figure 1S: Association between long-term HRV frequency-domain Z-score and aortic (n= 3154) and carotid (n= 1653) stiffness stratified by glucose percentiles in subpopulation without type 2 diabetes

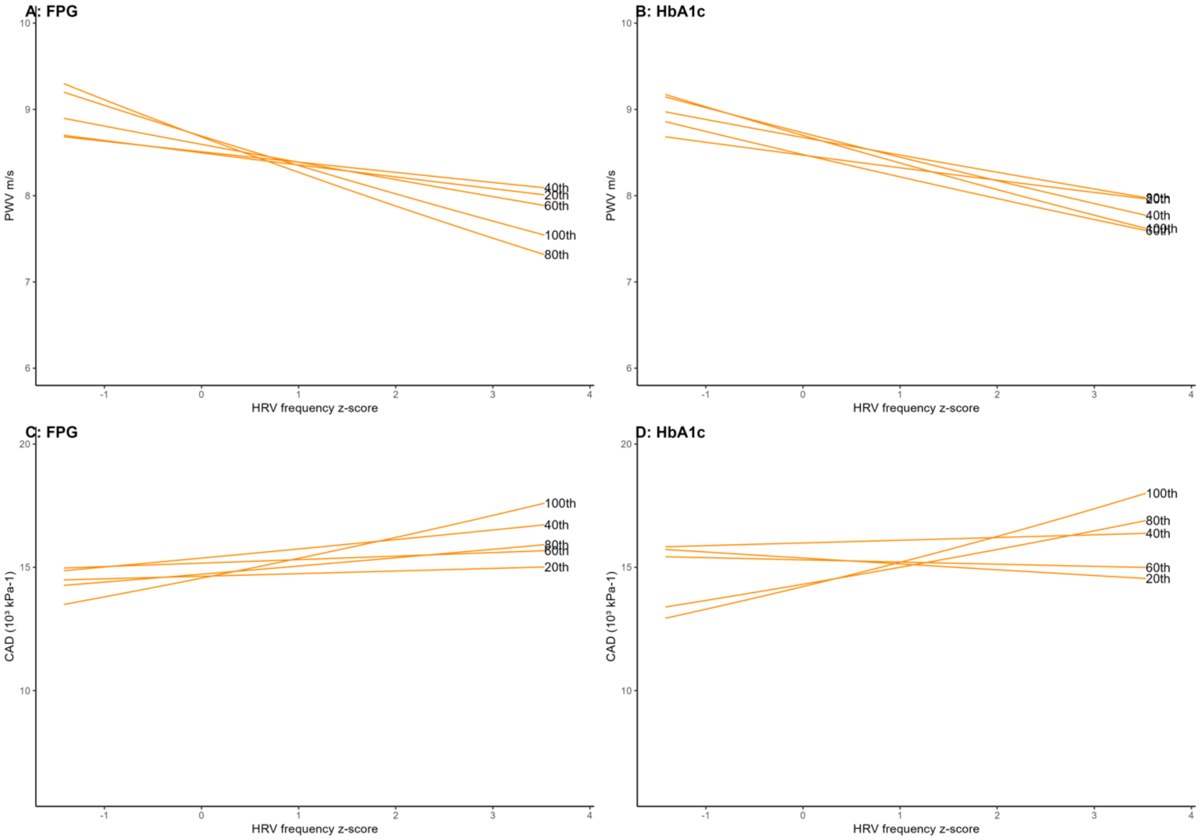

*Adjusted for age, sex, and mean arterial pressure*

### Figure 2S: Association between long-term time-domain Z-score and aortic (n= 3154 and carotid (n= 1653) stiffness stratified by glucose percentiles in subpopulation without type 2 diabetes

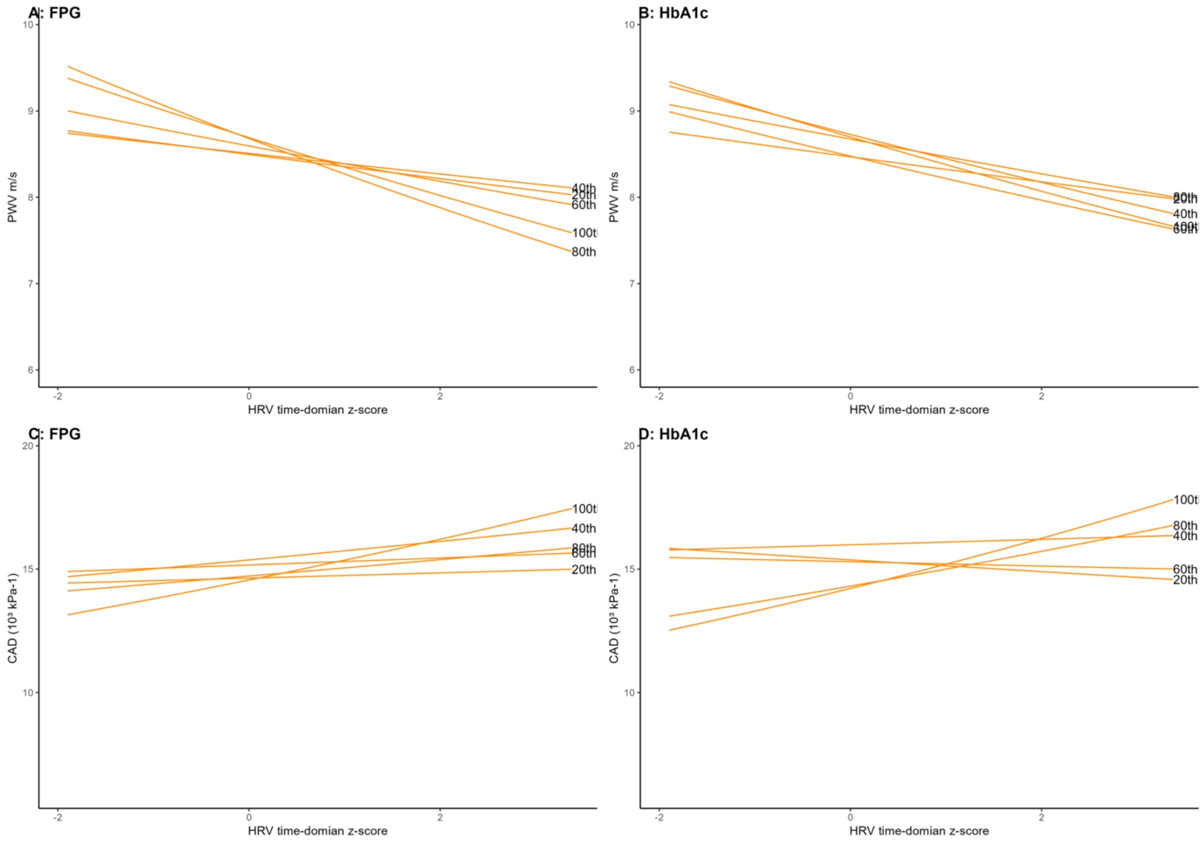

*Adjusted for age, sex, and mean arterial pressure*

#### Figure 3S: Association between long-term standardized HRV and arterial stiffness modified by diabetes status without users of beta-blockers

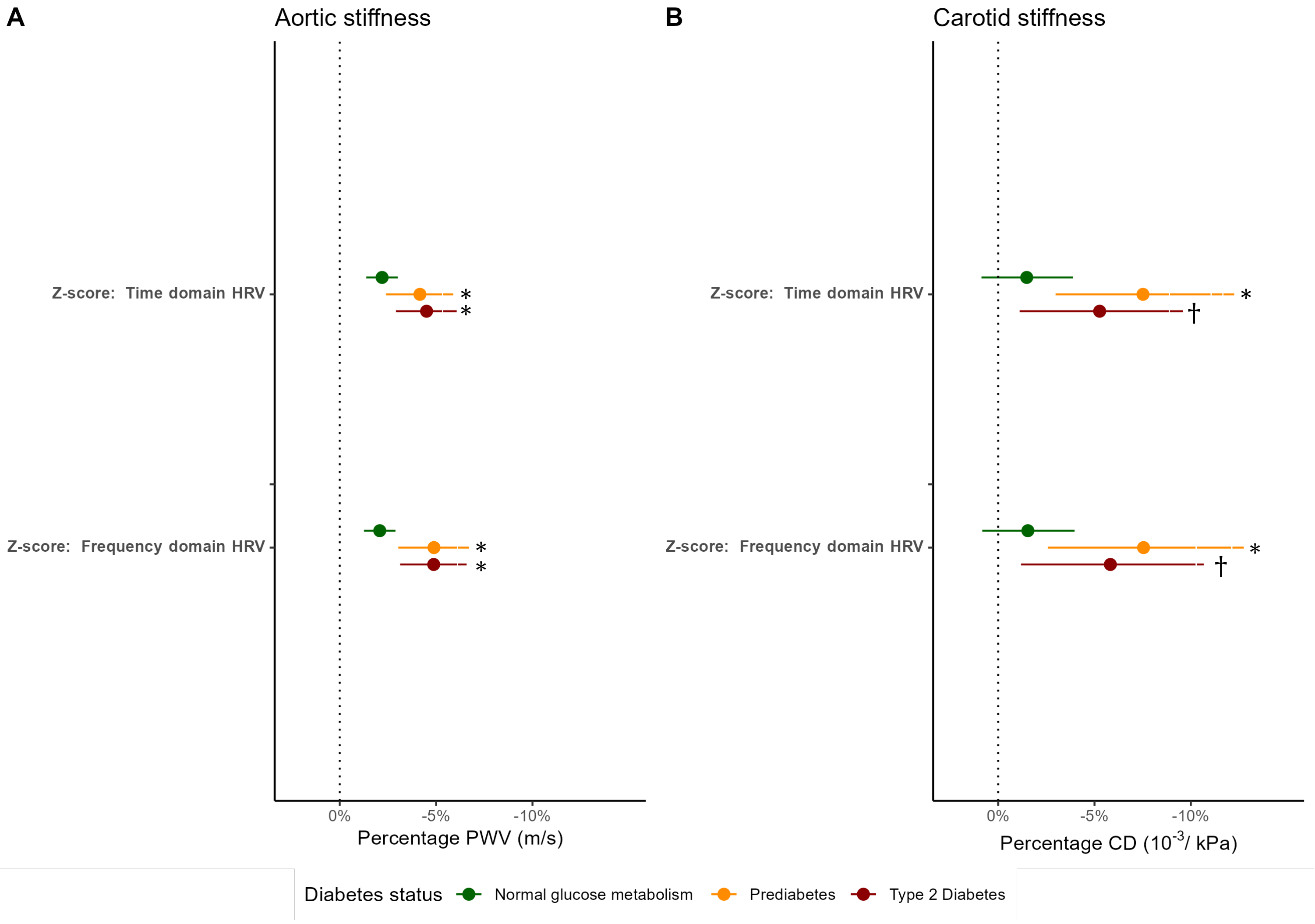

***A:*** *Percentage PWV per SD in time-domain and frequency-domain composite z-score by diabetes status* ***B:*** *Percentage CD per SD in time-domain and frequency-domain composite z-score by diabetes status. Estimates are adjusted for sex, age, educational status, mean arterial pressure, physical activity, smoking behaviour, alcohol use, body mass index, HbA1c, triglycerides, total-to-high density lipoprotein cholesterol ratio, lipid-modifying- and antihypertensive medication. Normal glucose metabolism was defined as reference group.*

**Interaction term p-value < 0.05*

*†Interaction term p-value < 0.10*
